# Open ecological, genomic and digital behavioural data resolve the 2026 MV Hondius Andes virus outbreak to the southern-Andean reservoir zone

**DOI:** 10.64898/2026.09.28.26364179

**Authors:** D.W. Redding, G. Albaladejo-Robles, J. G. Nino Barreat, D. Simons, J.L. Abbate

## Abstract

The 2026 Andes virus (ANDV) outbreak aboard the MV Hondius cruise ship resulted in 11 confirmed cases and 3 deaths and raised questions regarding the origin of infection, with much media focus resting on a landfill in Tierra del Fuego. We evaluated competing exposure hypotheses by assembling open data on reservoir host occurrences, rodent serosurveys, and historical ANDV outbreaks along with the index case’s self-documented, publicly-posted citizen-science digital record of their movements. Historical outbreaks associated with human-to-human transmission were concentrated within a restricted region spanning southern Chile and adjacent western Argentina, coinciding with the distribution of the principal reservoir host, *Oligoryzomys longicaudatus*, and areas identified as elevated risk by multiple ecological and genetic models. The index case was documented within this transmission focus shortly before the putative exposure period. Together, our open data approach was able to rapidly locate initial exposure within the established ANDV transmission focus and develop scenarios to prioritise further epidemiological investigation.

## Introduction

Andes orthohantavirus (ANDV, *Orthohantavirus andesense*) is a member of a diverse complex of genetically distinct, largely reservoir-specific orthohantavirus strains distributed across South America. It is a single-stranded, negative-sense RNA virus with a tripartite genome and is the principal agent of hantavirus pulmonary syndrome (HPS), a severe cardiopulmonary illness with high case-fatality (21.4-35.9%, Vial et al. 2023) Unusually among hantaviruses, ANDV is transmitted person-to-person via respiratory droplets, creating secondary transmission risk in healthcare and household settings, including documented clusters in Chile and Argentina (Wells et al. 1997, Padula et al. 1998, Martínez-Valdebenito et al. 2014, Martínez et al. 2020). The human-transmissible ANDV strain is maintained specifically by the long-tailed colilargo, *Oligoryzomys longicaudatus* with human spillover concentrated in the temperate forest and shrubland habitats of the southern Andes of Chile and Argentina (Padula et al. 2000, Palma et al. 2005). Experimental infection studies demonstrate efficient transmission in *O. longicaudatus* and, although spillover to *Abrothrix olivaceus* occurred experimentally, sustained onward transmission has not been shown in other rodent species, supporting a dominant maintenance role for *O. longicaudatus* (Padula et al. 2004).

On 3 April 2026, a previously healthy adult male (Patient 0) developed symptoms consistent with ANDV infection while aboard the MV Hondius cruise ship at sea (Andes Virus Outbreak Working Group,2026). Patient 0 had travelled through ANDV endemic regions of South America between November 2025 and March 2026, during which he publicly posted precise time and location of bird sightings to a citizen science website (https://ebird.org/). Detailed epidemiological investigation, including serological testing and clinical specimens, confirmed ANDV infection, and rapid sequencing associated the outbreak genetic sequences with those from previous outbreaks in Argentina in 2018/9 (Alpanez (2026, May 11), Gonzalez (2026, May 11), ECDC (2026, May 12), WHO (2026, May 13)). However, soon after the outbreak on the cruise ship was announced, authorities in Ushuaia, Tierra del Fuego, Argentina, were reported to be investigating a landfill near its point of departure (Risemberg et al. (2026, May 6), Zibell (2026, May 10)).

Human-to-human transmission of ANDV infection has not been reported from spillovers occurring outside the Andean mountains of southern Chile and Argentina (Bellomo et al. 2023). Despite extensive work examining ANDV phylogeography (Lopez et al. 1997, Padula et al. 2000, Palma et al. 2005, Bellomo et al. 2023) currently we do not yet know how the genetic structure and host specificity of ANDV varies quantitatively over space. Most importantly, we do not know how the travel trajectory of Patient 0 interacted with endemic risk areas in the weeks prior to symptom onset.

Here, we triangulate the spatial risk of Andes virus (ANDV) exposure from three independent lines of evidence: rodent seroprevalence surveys, the genetic relatedness of all available geolocated ANDV sequences, and the modelled distribution of candidate host species. We then overlay the reconstructed journey of the index case (Patient 0) onto the resulting risk surfaces to resolve where and when exposure to an infected reservoir most plausibly occurred. In doing this, we are asking a deliberately demonstrating our ability to ask an operational question: using only data already in the public domain at the time of the outbreak e.g. archived sequences, rodent serosurveys, reservoir occurrence records and the patient’s own publish digital behaviour footprint, can the likely site and timing of exposure be resolved rapidly and robustly enough to inform an active response? Our aim here is not a definitive account of ANDV diversity but a demonstration that pre-existing, openly available data can produce decision-adequate outcomes during a zoonotic disease outbreak.

## Results

The Andes orthohantavirus complex resolved into eight well-supported phylogeographic clades (Fig. 1, Extended Data Fig. 1), each clade largely restricted to a single reservoir rodent and bounded by that host’s geographic range. Whereas prior genomic work on this system characterised within-lineage markers of virulence and transmissibility (Bellomo et al. 2023), these clades represent the most parsimonous phylogeographic structure of the host-virus complex. A single, maximally supported monophyletic clade, (ultrafast bootstrap support = 100%, segments S/M), contained the pre-2026 human transmissible strains (Extended Data Fig. 1 – black stars; El Bolsón, 2014; Epuyén, 2018–19) and was overwhelmingly composed of isolates from *O. longicaudatus* (Fig 1b). In this clade, the M segment (124 tips) solely comprises *O. longicaudatus* and human exposures; in the taxonomically more widely-sampled S segment, of the 105 tips, there were three sequences from co-occurring native rodents (*Abrothrix longipilis* - 2, *Loxodontomys micropus* - 1) and two from the peridomestic, invasive *Rattus rattus*. We refer to this clade as the Andes virus strain clade and center the remaining analyses on it.

**Fig. 1.**
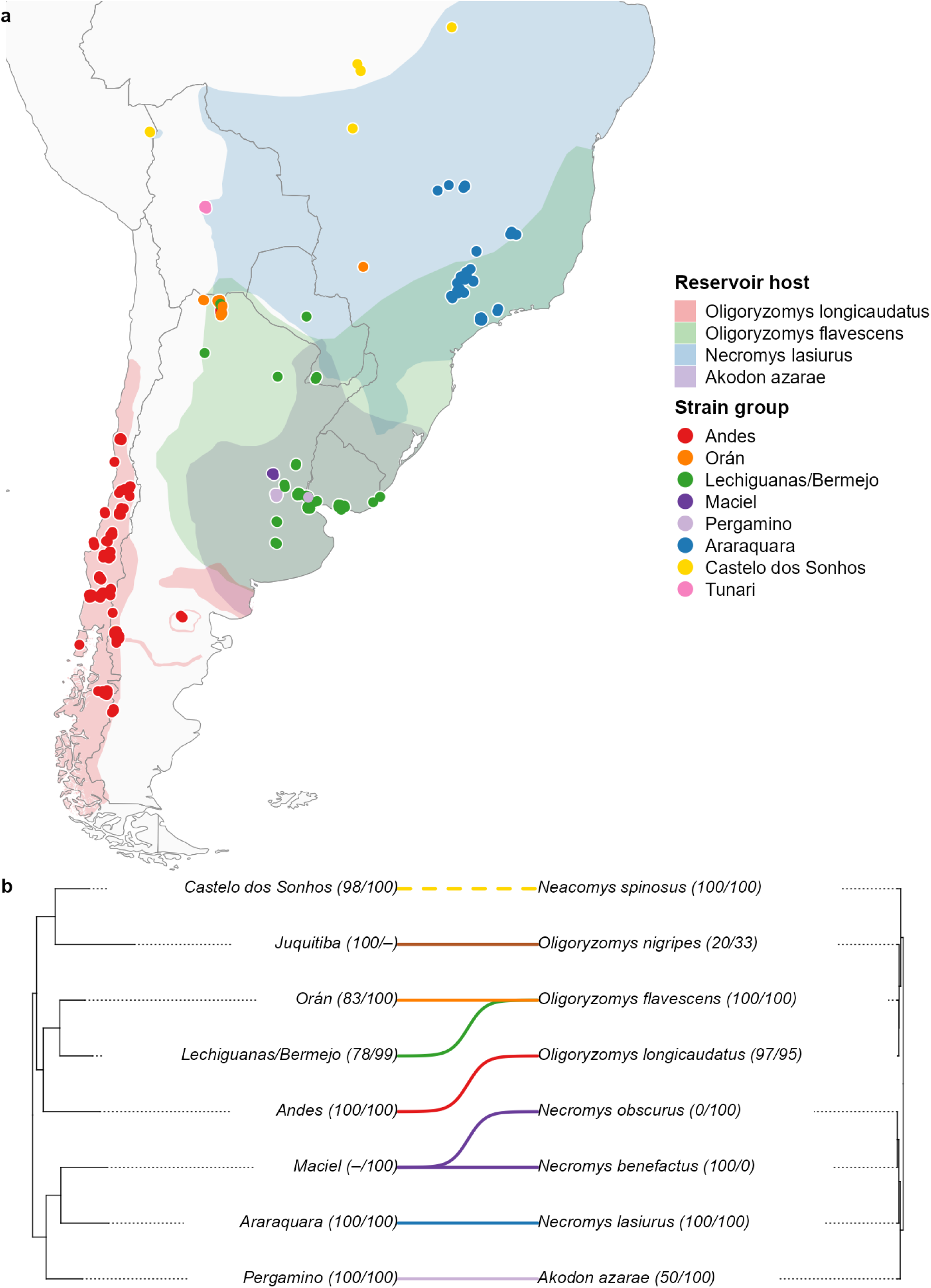
Andes orthohantavirus strains and their rodent reservoirs. (a) Geolocated sequences of Andes orthohantavirus (n = 387; S-and M-segment sequences combined, one point per accession) coloured by tree-defined strain group, over the shaded IUCN range polygons of the reservoir host of each well-supported strain. Maciel, Juquitiba and Castelo dos Sonhos, Tunari are plotted as points, but their host ranges are not shaded due to the reservoir being unresolved. (b) Tanglegram linking each strain (left; a single medoid S-segment representative) to its reservoir host (right; pruned mammal phylogeny). Strain tip labels give ultrafast-bootstrap support for that clade (Segment S/Segment M); host tip labels give host fidelity as (S/M), the percentage of that host species’ sequences assigned to the paired strain in the S and M segments, respectively.

We were able to geolocate 89% of the ANDV sequences and using a Bayesian isolation-by-distance model with a fixed effect for segment and random effect controlling for study, show that the genetic distance to the ANDV cluster increased steeply with geographic separation (n = 336, slope 0.036 per ln-km, 95% credible interval 0.034–0.038; R² = 0.76; p < 0.001; Fig. 2). This slope means, for example, a sequence sampled around 1,000 km away is roughly 17 percentage points more divergent than one 10 km away, up to around 20 percentage points divergence across the full sampled range. This slope value was stable irrespective of overlap and minimum length criteria (n = 217–357, slope = 0.036–0.037 per ln-km, R² = 0.75–0.79, all p < 0.001; Extended data Fig. 3). Matrix permutation supported this relationship with a positive correlation between geographic and sequence distance (Mantel S r=0.58 & M r=0.72, both p=0.0001). All the assigned sequences in the phylogeographic group containing the human-transmissible lineages were Andes virus strain clade and other strains formed more distant groups (Fig 2.).

**Fig. 2.**
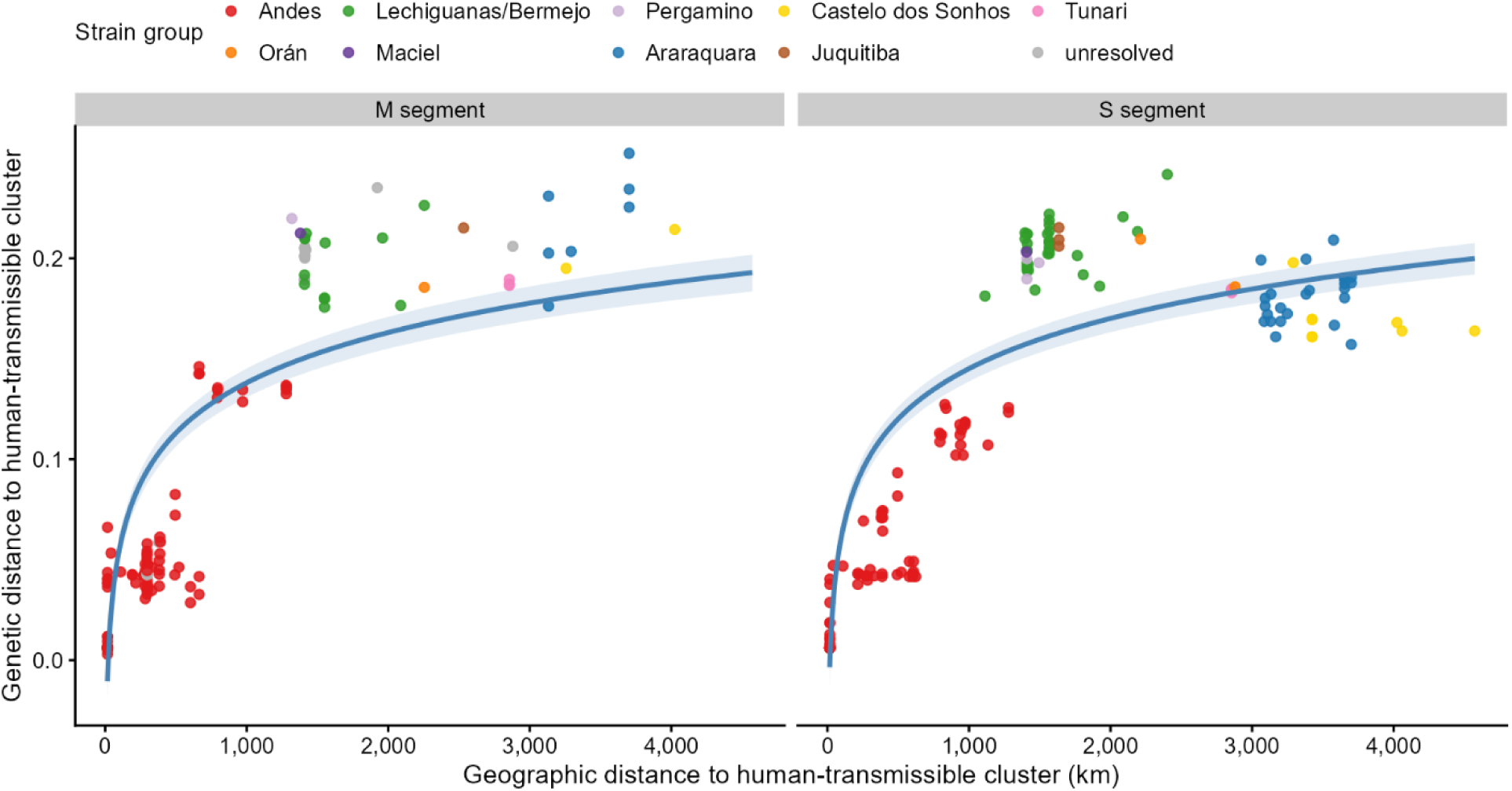
Isolation by distance in the Andes virus lineage. Great-circle geographic distance, per sequence, versus raw genetic distance to the human-transmissible ANDV, coloured by strain group and faceted by segment; line and band show the population-level fit and 95% credible interval of the Bayesian (INLA) isolation-by-distance model. Genetic distance increased with ln(geographic distance) (slope 0.036, 95% CrI 0.034–0.038; R² ≈ 0.76).

When examining ANDV seroprevalence (n = 564 from 22 studies with 18,551 animals tested, odds ratio 3.1, 95% credible interval 2.2–4.5; Fig. 3a,b) *O. longicaudatus* carried roughly three times higher prevalence than other native species, while congeneric *Oligoryzomys* species were equally as seropositive as *O. longicaudatus*. However, Andes viruses sequenced from *O. longicaudatus* were markedly more closely related to the Andes virus strain clade than those from native rodents including other congenerics (one-sided Wilcoxon p<0.001 for both segments; Fig. 3c,d), meaning congeneric sequences resolve instead to co-circulating members, such as Lechiguanas virus strain. The same three syntopic non-*O. longicaudatus* species noted in the phylogenies (*Abrothrix longipilis*, *Loxodontomys micropus*, *Rattus rattus*) hosted some closely related Segment S sequences, despite very low general prevalence levels (Fig. 3). However, *O. longicaudatus* has solid evidence (i.e. many seropositive animals from a high number of individuals tested) of ANDV infection in 26 independent sites, non-*O. longicaudatus* species such as the three noted above only had solid evidence of infection in 2 sites and both these sites also contained solid evidence of infected *O. longicaudatus* (>2% prevalence, n = 73, Binomial p < 0.001; Extended Data Fig. 4b).

**Fig. 3.**
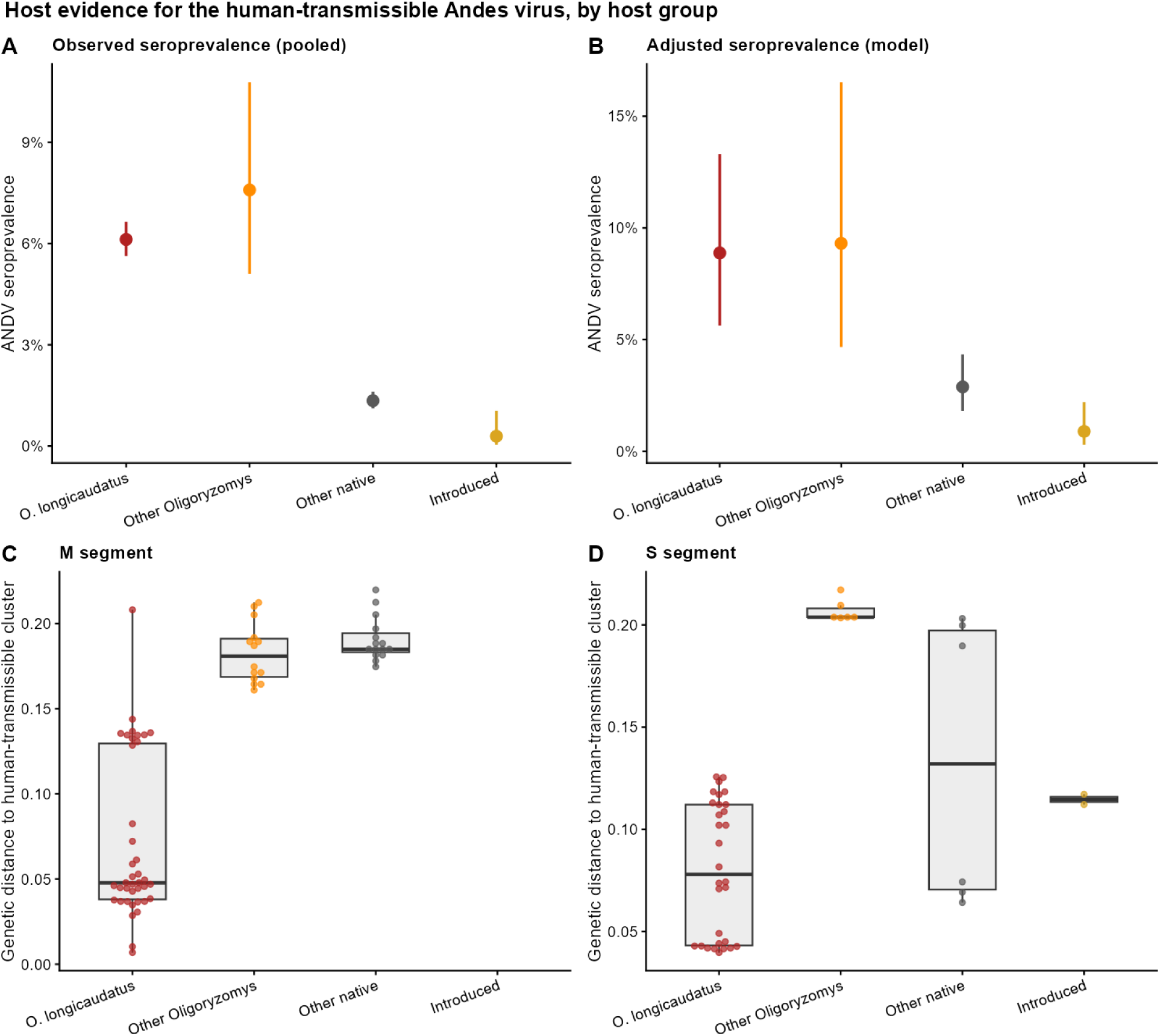
Andes virus seroprevalence rates among rodent groups and mean sequence distance to human transmissible strains. (a) observed and (b) Model-adjusted ANDV seroprevalence by host group (beta-binomial model, 564 records, 22 studies; IUCN range-cleaned). (c,d) Raw genetic distance to the ANDV cluster by host group for the M and S segments (box = IQR, points = sequences); *O. longicaudatus* is significantly closer than other native rodents (one-sided Wilcoxon rank-sum P <0.001 for both segments).

Focusing on *O. longicaudatus*, there were 1395 presence records on the global biodiversity information facility (GBIF, 2026) which when filtered resulted in 135 locations we used to build a MaxEnt model occurrence along with 10,000 background points, and 11 environmental predictors retained after collinearity filtering (from an initial 25). Of 50 candidate models (random feature/β-regularisation combinations), the 10 best by AICc were retained and averaged to final ensemble layer as all were well-calibrated, with continuous Boyce indices of 0.75–0.89 (Fig. 4a).

**Figure 4.**
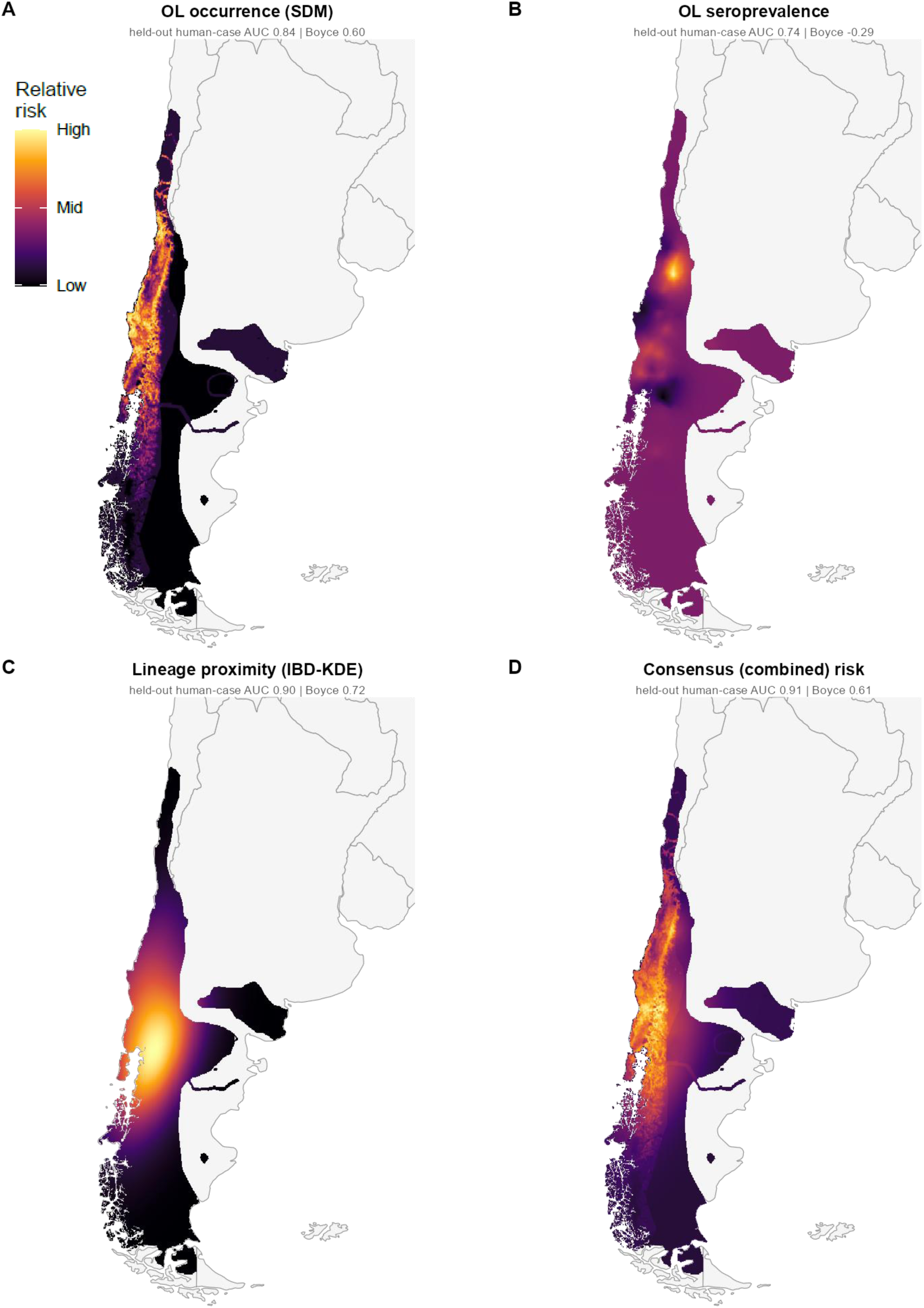
Three independent risk surfaces converge on north western Patagonia. (a) Reservoir occurrence (ensemble SDM), (b) reservoir seroprevalence (beta-binomial SPDE model), (c) proximity to the human-transmissible lineage (IBD-derived kernel surface), and (d) their consensus (per-cell mean). Surfaces are within-layer relative risk on a common grid, confined to the combined range of the three native potential hosts. Layers were weakly correlated (pairwise Pearson r = 0.03–0.45). Subtitles give discrimination of held-out ANDV human cases (area under the ROC curve; continuous Boyce index).

We then comparatively mapped spatial risk in southern South America of contacting ANDV using the models derived from the three independent datasets: the ensemble reservoir occurrence, a spatial prediction of ANDV seroprevalence, and Kernel density estimate (KDE) of the proximity to the human-transmissible lineage. While these layers showed only weak to moderate correlation (pairwise r = 0.03, 0.08 and 0.45) they converged on the same Andean–Patagonian strip of southern Chile and western Argentina (Fig. 4a–c). Validated against held-out ANDV human geolocated sequences (n = 92), all layers showed better than expected predictive ability (AUC = 0.74–0.90) though for the sequence-based layer this was partially confounded as the model likely included rodent sequences from the same locations. The consensus of the three layers, however, predicted case localities better than any individual layer (AUC = 0.92; Fig. 4d).

Finally, we reconstructed a detailed journey of the presumed index case (Patient 0) from their publicly submitted bird sightings and announcements about the patient’s movement from governments of the affected countries, resulting in an itinerary spanning four months with 57 geolocated time points (Extended Data Table 1). Overlaying the patient’s journey of onto our risk layers (Fig. 5) predicted exposure peaking twice in early-and late-January and again in mid-February while the patient was travelling through Patagonia and southern Chile. Human HPS cases in the ANDV endemic zone cluster in the summer–autumn months, when *O. longicaudatus* populations peak and rodent–human contact rises; the index case’s highest-risk window (January–February) falls within this seasonal peak (Martinez et al. 2010). Risk from all three layers fell to zero throughout much of the 2–6-week pre-symptomatic incubation period window, which the patient spent time in northern Argentina and Uruguay. We note a small reporting gap from 28th to 30th March when the patient was presumably in transit (Fig. 5). The highest-risk exposures appear to precede symptom onset (3 April) by approximately 7–10 weeks, at or beyond the outer limit of the recognised ANDV incubation period (7–39 days, median 18; Vial et al. 2006).

**Fig. 5.**
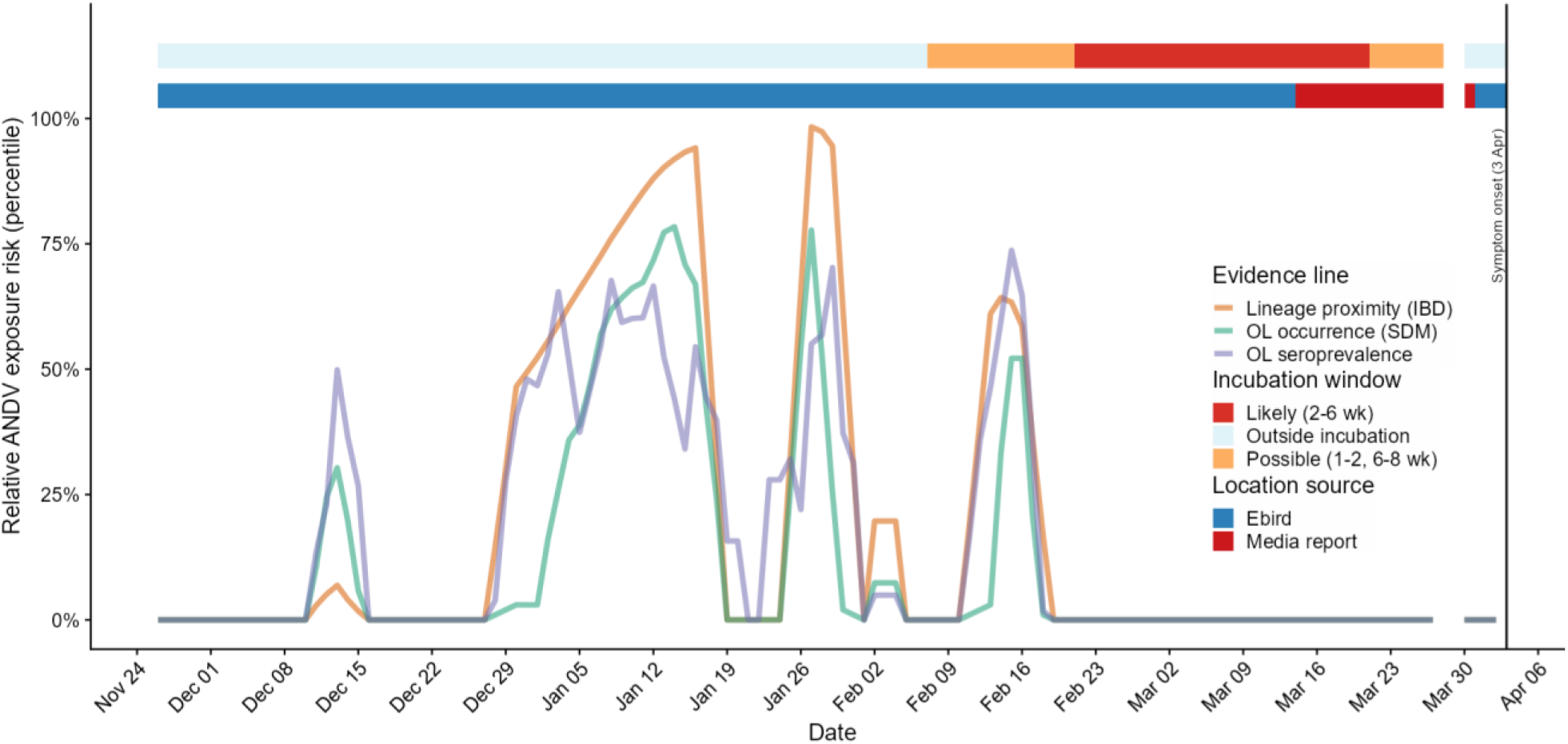
Index-case exposure-risk timeline. Daily relative risk (within-layer percentile, daily values smooth over a 3-day window) for the three layers along the index case’s reconstructed journey. The upper bars show the information source used to place the subject (eBird vs media report; white/orange/red) and the incubation classification relative to symptom onset (3 April 2026; blue/red).

## Discussion

The multimodal ecological, epidemiological and genomic datasets analysed here all place the likely origin of the 2026 ANDV outbreak strain in south-central Chile or across the border into western Patagonian Argentina, a region with well-documented ANDV endemicity and confirmed historical spillover events with onward human-to-human transmission. Temporally there is some inconsistency with established understanding, however, as the gap of 7 to 10 weeks is greater than the widely accepted ANDV incubation period (7–39 days, median 18; Vial et al. 2006). There are a set of non-mutually exclusive hypotheses that could explain this pattern:

1. Extended or heterogeneous incubation during zoonotic transmission. ANDV incubation shows documented variability, potentially modulated by host factors (viral load, immune kinetics, age, comorbidities; Vial et al. 2006, Martinez et al. 2020).
2. Broader reservoir range not captured in the current dataset could change the peak exposure date. As we note, members of the ANDV have been isolated from alternative rodent hosts in Argentina and Chile, and further isolations of human transmissible strains in other species could potentially extend the geographic footprint beyond *Oligoryzomys longicaudatus* distributions. However, our results support high maintenance host specificity (Extended Data Fig. 3d) in ANDV meaning this hypothesis remains highly unlikely. Consistent with this, effective reassortment appears rare in ANDV over macroevolutionary timescales (Rivero et al. 2026), making segment exchange a less likely route to rapid host-range expansion, with two caveats: First, reassortment detection is itself limited by some single segment sequencing, so an absence of observed reassortment does not establish that it is mechanistically impossible. Second, host-range shifts can in principle also arise through point mutation, albeit more slowly than through reassortment. The tight host specificity we observe is thus most parsimoniously explained by co-evolutionary constraint and the absence of detectable host-jumping rather than by an absolute inability to adapt; either way, a cryptic non-*Oligoryzomys longicaudatus* reservoir of the human-transmissible lineage remains unlikely on current evidence.
3. Exposure to an unidentified infected human. Without a confirmed rodent exposure event, we cannot rule out the possibility that Patient 0 contracted the disease from another human outside of the geographic range for zoonosis. However, historical human-to-human transmission of ANDV is characterised by subcritical chains (R0 rapidly falls <1) which typically generate distinct, highly visible case clusters. If Patient 0 had contracted the virus from a human source, that “true” index case would most likely have sparked a detectable cluster in Argentina or Chile. The apparent absence of such a local cluster points to Patient 0 as the primary zoonotic spillover event, rather than a secondary link in an extended human transmission chain.
4. Aerosolisation within a transiently contaminated microhabitat. With at least three reports mentioning that the patient travelled in a camper van (Centenera (2026, May 6), Horowitz et al. (2026, May 16), Kupferschmidt (2026, 11 June)), a persistent, breeding rodent infestation is not required to explain the infection. *O. longicaudatus* is an opportunistic forager (Piudo, 2011), overnight intrusion into a parked vehicle leaves infectious excreta within a poorly ventilated, enclosed space. Subsequent aerosolisation of this dried material, for example sweeping the vehicle weeks later, would trigger a high-dose exposure event. This mechanism could account for the extended 7-10 week gap between physical presence in the endemic zone and symptom onset, mirroring the established environmental exposure pathways of Sin Nombre virus (SNV) (Richardson, 2013).

This potential spatial and temporal decoupling of Patient 0’s physical presence in the endemic zone from subsequent symptom onset introduces further nuance to the surveillance of rodent borne diseases. If delayed aerosolisation within a transiently contaminated microhabitat represents a primary transmission mechanism, then this must be considered in future outbreak responses. For wilderness tourism, overland travel, and mobile lodging industries globally, this underscores a cryptic risk where a vehicle can act as a mobile incubator for infectious fomites long after exiting the high-hazard ecotone.

These findings illustrate both the power and limitations of phylogeographic approaches to outbreak investigation. Molecular evidence constrains geographic possibilities robustly but cannot always accurately resolve exposure timing or distinguish competing epidemiological scenarios with certainty. Retrospective collection of high-resolution travel data and rapid serological testing of contacts during early outbreak phases would substantially improve future attribution efforts. Notably, the entire assessment drew only on data already in the public domain and required no new field sampling or sequencing, so in principle it could be reproduced within the first days of an outbreak. Improving computation infrastructure for similar pathogen groups e.g. curated, analysis-ready reservoir-range, serosurvey and sequence resources (such as the ArHa database; Simons et al. 2025), and maintaining these resources between outbreaks alongside a pre-validated analytical pipeline, could allow triangulated exposure risk to be generated in near-real time.

A distinctive feature of our analyses is that the index case’s movements were recovered not from traditional epidemiological methods (e.g. interview) but from data the patient had published, in real time, to a citizen-science platform, containing both precisely dated and georeferenced observations. This points to a broader and still under-exploited opportunity in outbreak response: many people now continuously self-document their behaviour digitally including their locations and activities across citizen-science projects, social media and location-aware applications, generating fine-grained behavioural traces that are frequently available immediately and at a resolution formal surveillance has rarely matched. Such passively generated data can, in principle, uncover exposure-relevant data before official notification, for example early, geolocated social-media activity, for example, might have sharpened situational awareness in the opening weeks of the COVID-19 pandemic, when confirmed case data lagged the epidemic. Realising this potential responsibly requires explicit attention to consent and re-identification risk. While the locational data here were self-published to a public platform, harvesting private individuals’ data at scale raises materially greater ethical and legal obligations, as well as highlighting the non-representativeness of who self-documents and to data veracity. Handled within an appropriate governance framework, self-documented digital traces could become a routine, real-time complement to conventional outbreak investigation.

Each surface we present here is a statement of risk given the available evidence, and each inherits the structural biases of its source data (Simons et al. 2026). Critically, however, these three data streams carry different sets of biases e.g. literature-driven sequencing, opportunistic serosurveys and occurrence records, but converge on the same region and, in combination, discriminate fully held-out human cases better than any layer alone (AUC 0.92; Fig. 4d). A pattern generated purely by where sampling has been concentrated are unlikely to produce independently biased layers that both agree with one another and predict cases withheld from all of them. The value of the approach is thus not that any single dataset is complete, but that openly available, independently biased data can triangulate to a decision-adequate answer. Our *O. longicaudatus* suitability surface is concordant with previously published orthohantavirus and reservoir distribution models for the region (e.g. Andreo et al. 2011, Astorga et al. 2018), which likewise place the core suitable zone in the same Andean–Patagonian temperate forest–steppe. Indeed, this highlights that having expert-derived suitability layers in an epidemiological repository would remove the need for rapid reidentification of spatial risk for key host species, such as the one presented here.

Reinforcement biases, for instance, focusing efforts looking for a particular strain repeatedly in the same presumed reservoirs, will be amplified in synthetic analyses of publicly available data, such as the one we present here. We therefore looked to sensitivity-test our results throughout. For example, the exposure boundary is set by the reservoir’s independently-mapped range rather than by sequencing density, and the isolation-by-distance signal is invariant to sequence length, coverage filtering, genome segment and to dropping individual reference anchors (Extended Data Fig. 3). Additional surveys could indeed refine fine local detail but are unlikely to relocate a boundary fixed by host range and corroborated across independent data streams. However, the phylogeographic structure of the ANDV clade means exposure to human transmissible strains appears tightly geographically focused and located on one side of an imposing geographical barrier. With climate change and ongoing transportation infrastructure development potentially altering the permeability of this barrier, further increasing resources to aid efforts already occurring within the affected countries to understand processes of infection and transmission in endemic foci remains essential given ANDV’s recurring and impactful spillover potential.

## Methods

Sequence data and inclusion. All available Andes virus S-and M-segment sequences (n = 739) and their metadata were obtained from NCBI Virus (NCBI Virus). Each sequence’s geographic origin was resolved by taking the most authoritative coordinate available: literature-derived or manually corrected localities, then curated coordinates from the ArHa hantavirus database (Simons et al. 2025), and remaining data points with GenBank localities were geocoded with the OpenStreetMap Nominatim API (nominatim.org). Those with just country data were left without coordinates. Data-restricted accessions, vaccine candidates, 49 vesicular-stomatitis-virus pseudotype constructs of strain Chile-9717869, the laboratory host *Mesocricetus auratus*, two non-reservoir bats (*Carollia perspicillata*, *Desmodus rotundus*), strains designated “hamster lab passages” and the L segment (excluded due to poor coverage) were removed. After cleaning, 560 sequences remained (477 in the aligned S/M phylogenetic set: S, n = 217; M, n = 260), of which 498 (≈89%) had usable coordinates (Supplementary Table 1). Sequence lengths were heterogeneous, spanning short diagnostic fragments to complete segments (median 936 bp for S and 473 bp for M; range 150–3,696 bp), reflecting three decades of accumulation under differing sequencing aims. Rather than discard the many short records, all pairwise genetic distances were computed only over the region two sequences actually share (overlap masking; see Isolation by distance), and the robustness of every distance-based result to this length heterogeneity is demonstrated explicitly (Extended Data Fig. 3a).

Phylogenetics and clade assignment. Segments were aligned separately in MAFFT (version 7, Auto strategy, Katoh & Stanley 2013) and the alignments manually cleaned against a per-segment reference. Maximum-likelihood trees were inferred in IQ-TREE (multicore version 1.6.11, Nguyen et al. 2015; ultrafast bootstrap, Hoang et al. 2018) under the substitution model GTR+G with 4 rate categories, with branch support from SH-aLRT and 1,000 ultrafast-bootstrap replicates; trees were midpoint-rooted and, for display, nodes with ultrafast bootstrap < 50 collapsed. The two strains that did not form strongly supported monophyletic clades (Extended data Fig. 1) were those labelled Lechiguanas and Bermejo, and these sequences were then relabeled Lechiguanas/Bermejo. Using clade monophyly — a clade being a group of sequences descended from a single common ancestor — and the geographic ranges (IUCN, 2025) of the hosts we assigned each tip to a viral strain by its nearest reference tip in patristic distance, and flagged any record whose recorded host fell outside that host’s IUCN range at its stated locality (introduced commensals/peridomestics exempt) as a coordinate or host error. These flagged records were excluded from the distance-based analyses (isolation by distance and the kernel risk surfaces below), such that 336 sequences were included in the isolation-by-distance model. Four records were flagged and removed on this basis: two *Oligoryzomys longicaudatus* sequences geolocated to Orán (∼2,000 km outside the species’ range), one *Necromys obscurus* record at Maciel, and one sequence lacking a resolvable locality. As expected for spatial-genetic data, model residuals retained the positive spatial autocorrelation that shared ancestry produces (Moran’s I = 0.54–0.57, P < 0.001) and the slope is confirmed both by a Mantel test on the full pairwise matrices and by a study-level random-effect model, and is stable to removing individual reference anchors (Extended Data Fig. 3). We also estimated the location of data from the seroprevalence dataset we used below for illustration (Extended Data Fig. 2)

Isolation by distance. For each retained sequence, we computed two weighted-mean summaries of distance to the human-transmissible anchor set: First, an overlap-masked raw genetic distance (normalised Hamming distance, with pairwise deletion of gaps and ambiguities). Second, a great-circle (Haversine) geographic distance weighting each anchor by a phylogenetic-redundancy weight (1.5 for the least-redundant anchors, otherwise 1.0) multiplied by the reciprocal of the number of anchors sharing its coordinate. A pairwise genetic distance was initially treated as missing unless the two sequences overlapped at more than max (0.25 × shorter non-gap length, 250 bp), and a sequence was retained only if it had at least 250 non-gap bases and at least a fraction f of its segment’s 90th-percentile non-gap length (f = 0.10 for the primary analysis). Genetic distance was modelled using the natural log of geographic distance with a segment term using a Gaussian INLA model; we report the pooled (fixed-effects-only) slope as the headline estimate, with a study-level random intercept fitted as a sensitivity giving a very similar slope. From this model we measured the posterior slope with 95% credible interval, a Bayesian R² (variance of the linear predictor divided by that variance plus the residual variance, from 2,000 posterior draws), cross-validatory probability-integral-transform calibration, and a Mantel test on the full pairwise matrices (9,999 permutations; residual missing genetic cells mean-imputed); the model’s slope stability while varying coverage fractions (f = 0.05–0.25), overlap and length thresholds (Extend Data Fig. 3). Because many sequences are short, this coverage-fraction sensitivity is the decisive check that the decay is not a fragment-length artefact: progressively restricting the set to longer sequences (f = 0.05 to 0.25) leaves the slope essentially unchanged (0.036–0.037 per ln-km; Extended Data Fig. 3a). To visualise the fitted decay, each sequence’s genetic distance was inverted through the fitted intercept and slope to the geographic distance the model implies and weighted inversely to it; a weighted two-dimensional kernel density (R package ks) over unique localities was spatially predicted on a raster and rescaled to 0–1. The kernel bandwidth was selected automatically using the plug-in estimator (Hpi; Wand & Jones 1994). Per-sequence genetic distance to the human-transmissible cluster was compared between host groups by one-sided Wilcoxon rank-sum tests. Given *O. longicaudatus* had by far the smallest genetic distance to the human-transmissible cluster, the reservoir and distribution analyses below therefore focus on this species. However, from the 229 combined S and M sequences *Abrothrix longipilis* (2) and *Loxodontomys micropus* (1) had sequences that were located in the same overall Andes virus strain clade, so they could not be completely discounted as being a spillover source at this stage.

Reservoir-host analyses. With the phylogeographic analysis supporting *O. longicaudatus* as the principal but not sole reservoir of the human-transmissible clade, we then quantified its ANDV seroprevalence relative to the other rodents with which it co-occurs. Rodent serosurvey records were obtained from the ArHa serodatabase (Simons et al. 2025) and records retained where the reported pathogen matched Andes virus in only the strict sense (one of "ANDV", "Andes virus", "Andes hantavirus", "Andes orthohantavirus", "*Orthohantavirus andesense*", and capitalisation variants), and the assay was serological rather than molecular, the count was valid (0 ≤ positives ≤ tested) and host species and coordinates were present and within their IUCN range — 564 records from 22 studies. No temporal window was applied relative to the 2026 outbreak; retained surveys span 1995– 2021 (concentrated in 2000–2019), and the host-group contrast is stable across an escalating series of controls including a study random effect, a seasonal term and a Matérn spatial field (Extended Data Fig. 4a). Due to the number of records, and in order to make reasonable group comparisons, hosts were grouped as either *O. longicaudatus*, other *Oligoryzomys*, introduced (*Rattus rattus*, R. norvegicus, *Mus musculus*), or other native rodents. The other-*Oligoryzomys* group comprised three congeners, with all records identified to species (*Oligoryzomys flavescens*, *Oligoryzomys nigripes* and *Oligoryzomys chacoensis*). Seropositive counts were modelled as Beta-Binomial by integrated nested Laplace approximation (INLA) using host group, assay group (ELISA, strip immunoassay or unspecified), log number tested (centered and scaled), a study-level random intercept, and a cyclic second-order random-walk seasonal term of sampling month. Fixed-effect priors were Normal(0, SD 1.5) for the intercept and Normal(0, SD 1) for coefficients, and random-effect precisions had penalised-complexity priors with P(σ > 1) = 0.5. From this model we measured host-group odds ratios versus other native rodents and posterior-predicted prevalences (using the reference assay, using mean effort, and with random effects set to zero), with the contrast confirmed across a series of controls (Extended Data Fig. 4a). To test whether non-reservoir rodents are infected independently of *O. longicaudatus*, records were aggregated to 0.1° sites within the three-species range; at sites where *O. longicaudatus* and at least one other native rodent were both tested, a species was scored as solidly infected where its Beta-Binomial (Jeffreys-prior) posterior placed prevalence credibly above a floor (P[prevalence > 2%] ≥ 0.95; and 0.5% and 5% for sensitivity), and the frequency with which *O. longicaudatus* versus a non-*O. longicaudatus* species was the sole infected species was compared with a binomial test on discordant sites (Extended Data Fig. 4b). We extrapolated a seroprevalence layer using the posterior mean of a Beta-Binomial Matérn spatial model fitted by the stochastic partial differential equation (SPDE) approach in a Lambert azimuthal equal-area projection (mesh maximum inner/outer edge 70/300 km, 25-km cutoff), with penalised-complexity priors of median range ≈ 150 km and P(σ > 1) = 0.01, projected to a 0.1° land grid.

### Species distribution model

Because the phylogeographic (isolation-by-distance) and serological analyses both identify *O. longicaudatus* as the likely sole reservoir of the human-transmissible lineage, we modelled the potential distribution of this species alone. For this, we used the presence-only Maximum Entropy modelling algorithm (MaxEnt) (Phillips et al.2006). After filtering and selection, we obtained 124 unique spatial presence records of *O. longicaudatus* from the Global Biodiversity Information Facility (GBIF, https://www.gbif.org/). In addition to these occurrence data, we generated 10,000 background points by sampling the study area using a density kernel derived from *O. longicaudatus* presence data (e.g Barbet-Massin et al., 2012; Jarnevich et al., 2017; Barber et al., 2022). Presence and background points were modelled against 25 different environmental predictors; land cover heterogeneity was included using the cropland, grassland, and tree cover extracted from the European Space Agency WorldCover dataset (Zanaga et al., 2021); Climatic information was retrieved from BIOCLIM Version 2.1 (Karger et al., 2017). This dataset contains 19 different climatic variables with ecological potential derived from long-term monthly time-series of temperature and rainfall, and topographic heterogeneity was included in the form of terrain slope, aspect, and ruggedness (Horn, 1981; Wilson et al., 2007). To calculate these variables, we used the elevation data derived from the Shuttle Radar Topography Mission (SRTM) (NASA JPL, 2013) and the GTOPO30 (USG,1996) (Fick and Hijmans, 2017).

After controlling for collinearity (Variation Inflation Factor lower than 5, O’ Brien, 2007) and correlation (with a threshold of 0.7), only 11 environmental predictors were used: cropland, grassland, tree cover, slope, aspect, and 6 climatic variables BIO - 2, 3, 8, 9, 15, and 18. Using this combination of presence/background records, a 70:30 (training: testing) split, and environmental predictors, we fitted 50 different MaxEnt models. Each model was the result of a random selection of fitting features and β-regularisation parameters (range 1-6) (Redding et al.,2017; Albaladejo-Robles et al., 2025). This way, we were able to generate a wider range of model responses and distribution scenarios.

From these models, we only selected the 10 best-performing ones. Models were first ranked according to their Akaike Information Criterion (AICc) (e.g Stoica and Selen, 2004). The first 10 best-fitted models were then evaluated using the continuous Boyce index (Boyce et al., 2002). We set a minimum threshold of 0.25 to accept or reject the models. As a result, all best-ranked models (AICc) were selected (Boyce index values between 0.75 and 0.89). Selected models were then projected across the study region and averaged to create the potential distribution of *O. longicaudatus* (Fig 4.a).

All spatial information was coerced to the same spatial resolution of 2.5 arc-min (∼4.6 km at the equator) and coordinate reference system (WGS84 EPSG: 4326) (Further details on data processing, model fit and selection can be found in the supplementary methods).

Risk surfaces, consensus and validation. The three related layers based on the independent datasets: reservoir occurrence (the SDM), reservoir seroprevalence surface, and proximity to the human-transmissible lineage (the kernel surface outlined above) were placed on a common 0.05° grid (bilinear resampling), confined to the union of the IUCN ranges of those species with known ANDV sequences: *O. longicaudatus*, *Abrothrix longipilis* and *Loxodontomys micropus*, and expressed as within-layer percentiles (positive cells ranked to 0–1 and true zeros set to 0), with layer similarity assessed by pairwise Pearson correlation. A fourth layer was created using their per-cell mean as a consensus and their per-cell standard deviation as a representation of the between-layer disagreement (Extended Data Fig. 5). For validation, every ANDV human-case locality (n = 92; Extended Data Fig. 5) was withheld and the lineage-proximity kernel surface refit with no human cases, so all the risk surfaces were built solely from host data; each layer and the consensus were then scored at all withheld case localities against 10,000 random background points by the area under the ROC curve and the continuous Boyce index (R package ecospat).

### Index-case trajectory

The itinerary was reconstructed from the travellers’ eBird checklists (Sullivan, 2009) and official media reports (coordinates randomly displaced for privacy), each waypoint carrying an information-source category. Same-date records were collapsed to a daily mean position and linearly interpolated between consecutive known waypoints; a single documented gap where the patient’s location was unknown (28–29 March) was left as a break. Each daily position was scored on the three percentile surfaces, with days inside the three-species range floored to a small value for visibility, and days off the range set to zero, and smoothed with a centered 3-day mean. We set the incubation windows using literature sources at: likely (2–4 weeks), possible (1-2 weeks and 4-6 weeks) and unlikely (before 1 week and after 6 weeks; Vial et al. 2006, Andrews & Bogoch 2026).

### Implementation

Automated workflows, isolation-by-distance regressions, and geospatial modeling pipelines were executed using R version 4.5.1. Large language model infrastructure (Claude Opus 4.8, Anthropic, San Francisco, CA) was used strictly as an auxiliary tool to optimize code architecture, enhance parallelized spatial handling formatting, and assist in the structural drafting of algorithmic code blocks and to create integrity tests. In accordance with institutional reporting standards, all final code outputs, statistical diagnostics, and model iterations were vetted, audited, and verified manually by the authors to guarantee computational integrity. R packages used: Bayesian models were fitted with R-INLA/fmesher; phylogenetics with ape, phytools and ggtree; kernel density with ks; the Mantel test with vegan; the continuous Boyce index with ecospat; spatial handling with sf, terra and rnaturalearth; and great-circle distances with geosphere (for package versions see supplementary methods); MaxEnt models were fitted with using the 3.4.3 version using the interface built within the dismo R package version 1.3-14. Random seeds were fixed throughout for posterior sampling, the validation hold-out and kernel resampling.

## Acknowledgments

eBird data were made available through the Cornell Lab of Ornithology, and we thank M. Iliff and C. Wood for validating dates and locations against data not publicly shared.

## Author Contributions

D.W.R. designed the study, gathered the data, conducted the risk analyses, interpreted results and co-wrote the manuscript. J.L.A. coordinated data harmonization, validated patient location and epidemiological data, conducted the risk-by-patient location analysis,and co-wrote the manuscript. G.A.R. built the species distribution models and J.G.N.B. extracted localities, hosts and sources from the GenBank data. D.S. provided data and codesigned the analysis for the seroprevalence data. All authors reviewed, edited and approved the final manuscript.

## Data Availability

Code for phylogeographic analysis (isolation-by-distance regression, kernel density estimation, and ecological occupancy modelling) is available at https://github.com/BioDivHealth/ANDV_MVHONDIUS. All data necessary to reproduce the analyses are available in the supplementary materials or from the corresponding author upon request. The eBird trajectory data shared have been treated with random displacement of positions for privacy purposes. All risk maps are available and explorable with open access at [https://www.epiwise.com]. IUCN range polygons are not redistributable.

## Ethics Statement

This analysis is based on publicly available sequence data and de-identified epidemiological case information. No direct human or animal studies were conducted.

## Supplementary Methods - Species Distribution Modelling

### SDM data filtering and processing

To collect presence data for *Oligoryzomys longicaudatus* (*O. longicaudatus*) first we performed a taxonomic revision, gathering all the available information from the IUCN database and the Integrated Taxonomic Information System (ITIS) (both consulted in May 2026) (https://www.itis.gov/). These online repositories were used to extract all the upstream taxonomic information of the species and to gather synonym information for our target species. This was necessary in order to gather species presence records from the Global Biodiversity Information Facility (GBIF, https://www.gbif.org/). GBIF species presence records are composed of collections of datasets with different formats and quality. Data sources varied from citizen science observations to official surveys (Hughes et al., 2021). This can create taxonomic mismatches between different datasets within GBIF. Similarly, the IUCN Red-List follows its own taxonomic standards that might not match those of other big datasets (IUCN 2024). For this reason, a taxonomic matching was necessary to efficiently combine the spatial data from the IUCN and GBIF, and to gather all the spatial data for the species. These databases were accessed using the *rredlist* R package version 1.0.0 (Gearty and Chamberlain, 2025) and the *taxize* R-package version 0.9.100 (Chamberlain et al., 2013). GBIF also offers complete taxonomic information for the species, which can be accessed through taxize. However, previous experience has shown that erroneous metadata and synonym associations make it less reliable than those from ITIS or the IUCN, so all taxonomic information was adapted to follow the IUCN standards and format (IUCN, 2024).

Presence records were downloaded from GBIF using the rgbif R package version 3.8.1 (Chamberlain et al., 2025) using the IUCN species binomial names along with all their possible synonyms. In total, we were able to retrieve 1,395 *O. longicaudatus* records (human observations only) for the 1970-2025 period (GBIF.org, 12 May 2026). To clean these spatial records, we ran a hierarchical filtering approach. First, we excluded all the records that, according to GBIF data standards, contained inaccurate data (e.g coordinates derived from regional or national polygons) or incomplete metadata (e.g country derived from coordinates). This filtering was applied in the GBIF query. Second, we used the CoordinateCleaner R package version 2.0-20 (Zizka et al., 2019) to exclude points with erroneous/conflicted metadata, erroneous coordinates, or that corresponded to museums and other institutions known for hosting specimens’ collections. Third, because some GBIF data are not professionally collected, we used the species’ IUCN range information to remove observations outside the areas where the species has been described as present. In all cases we added a buffer of 1 arc-degree (∼100 km at the equator) to this range information to account for potential spatial bias. Finally, during the modelling phase, all points with duplicated coordinates or no environmental variable coverage were removed. As a result of this filtering, we obtained 135 unique spatial records of *O. longicaudatus* for modelling its potential distribution.

### Environmental predictors

To model the potential distribution of *O. longicaudatus*, we selected a combination of climatic, land-cover composition, and topographic variables. In the case of the climatic variables, we used the BIOCLIM dataset Version 2.1 (Karger et al. 2017) (https://chelsa-climate.org/bioclim/). BIOCLIM contains 19 different climatic variables (Table S3) with ecological potential derived from long-term monthly time series of temperature and rainfall. Due to its resolution, spatial cover, and diversity, BIOCLIM is frequently used in ecological studies, niche analysis, and species distribution modelling (e.g. Beaumont et al., 2005; Booth et al., 2014; Albaladejo-Robles et al., 2025). This information was retrieved at a 2.5 arc-minute resolution (∼ 4.6 km at the equator).

The 19 standard BIOCLIM (bioclimatic) variables used in WorldClim, ANUCLIM, CHELSA, and most species distribution modelling workflows. These variables are derived from monthly temperature and precipitation data and summarize annual trends, seasonality, and climatic extremes, as follows:

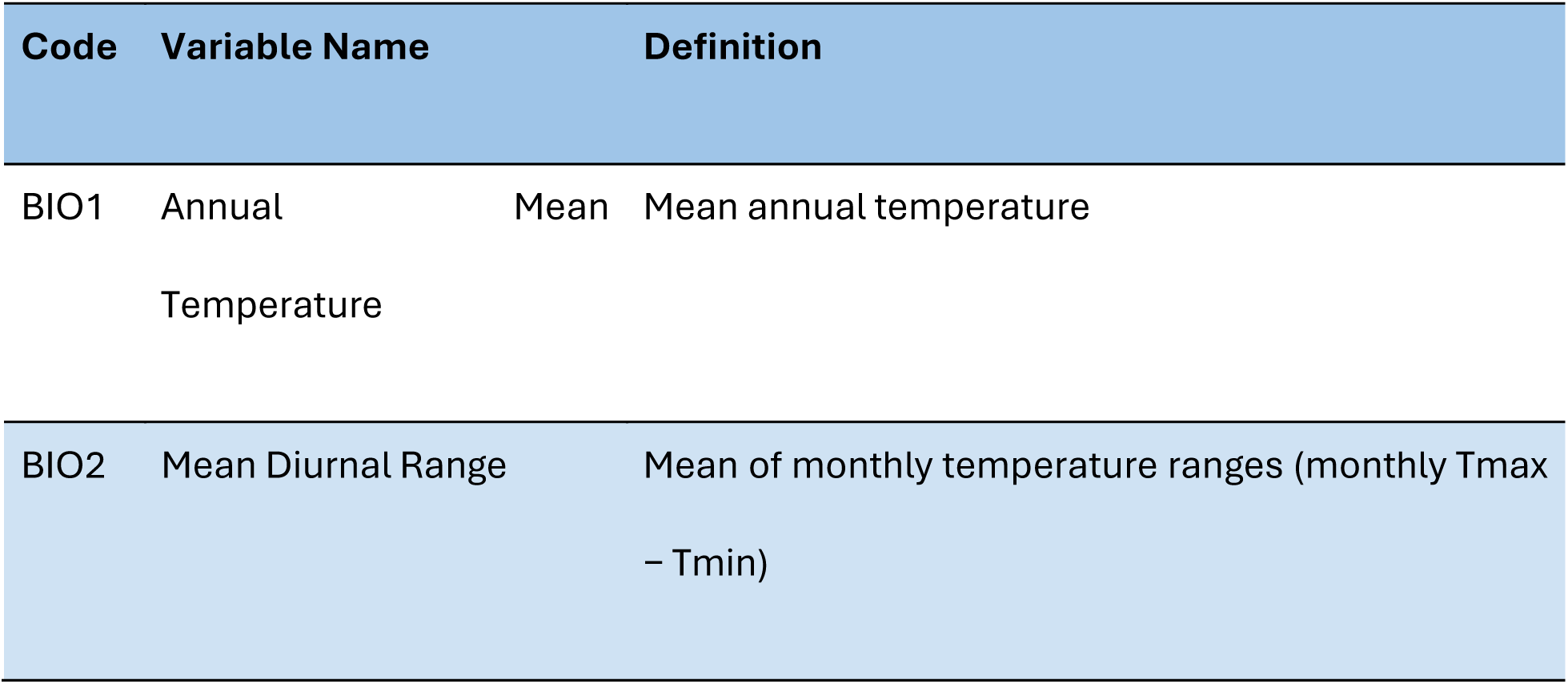
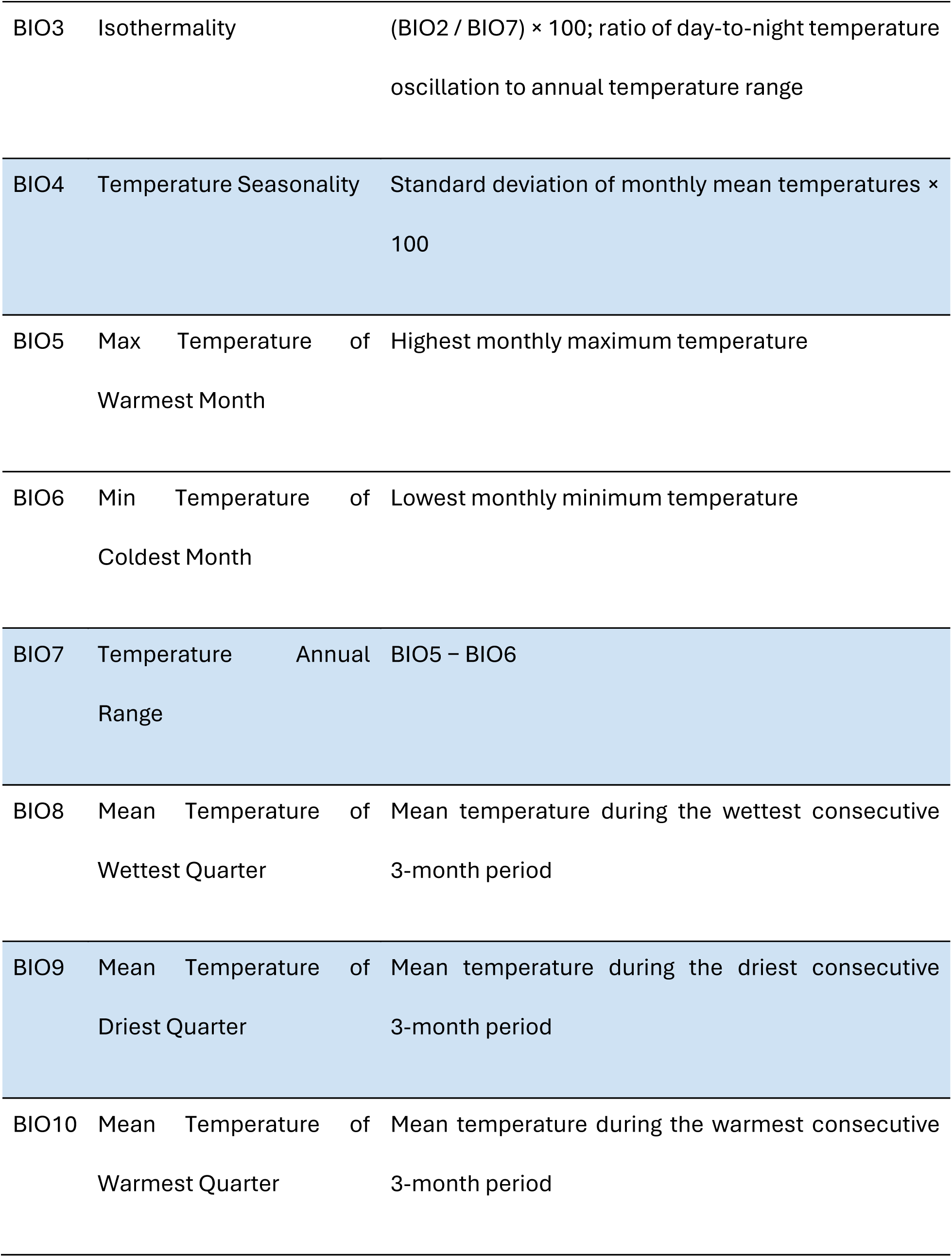
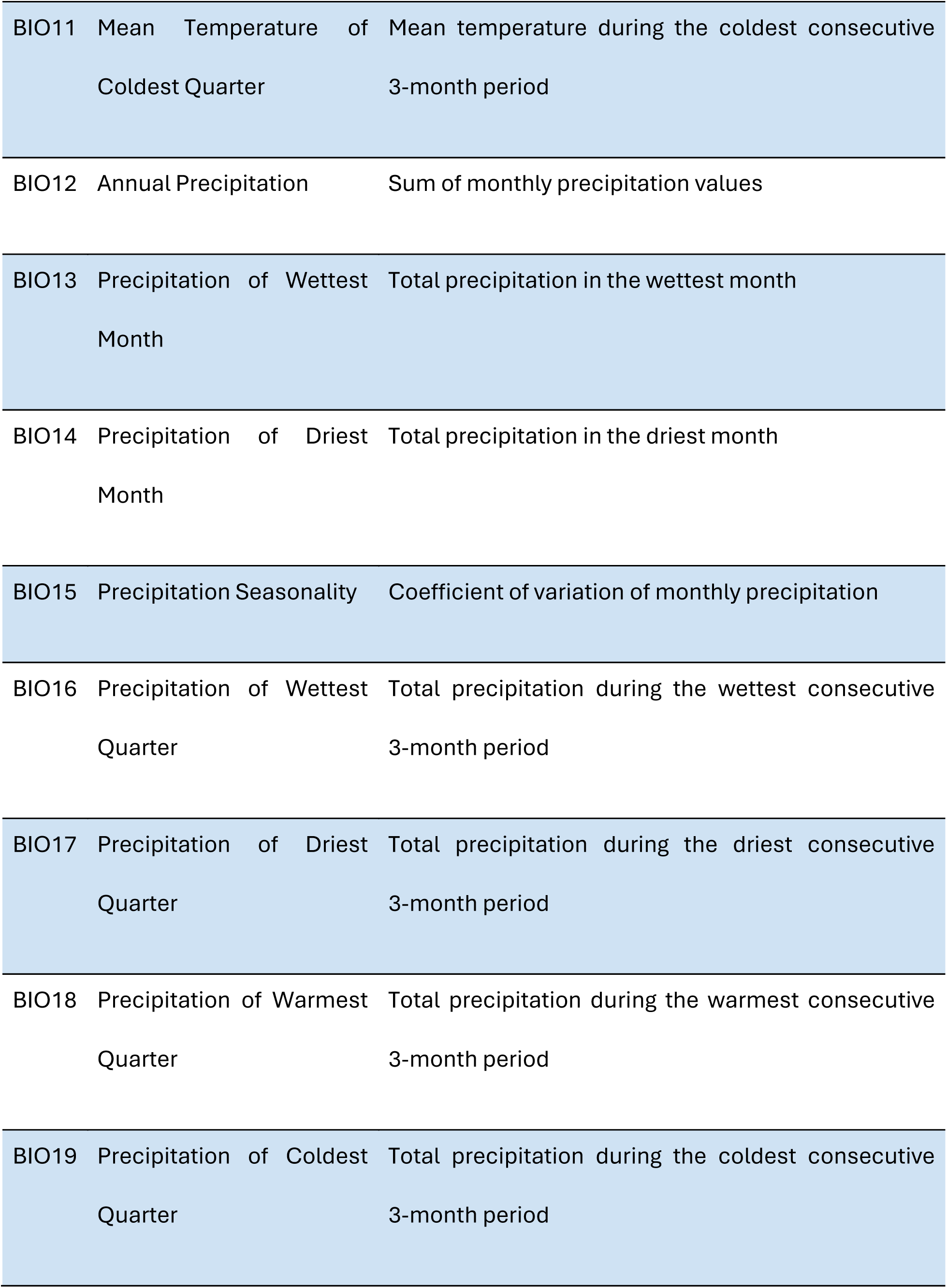

In addition to this information, we obtained land-cover composition information from the European Space Agency WorldCover dataset (Zanaga et al., 2021, https://esa-worldcover.org/en). This dataset contains a global land-cover map, at a maximum resolution of 10 metres, for 11 different land-cover classes in concordance with the UN-FAO’s land Cover Classification (Di Gregorio and Jansen, 2000). For the modelling of the potential distribution of our target species, we selected three land-cover types: tree cover, grassland, and cropland. These variables cover a wide range of land-use types and summarise the structure and composition of the landscape (e.g, Li et al., 2017). Variables are presented in the form of rasterised global maps on which each pixel reflects the percentage of total cover, between 0 and 1, of a given land-cover type (Zanaga et al., 2021). This data was obtained at a 30 arc-second resolution (∼1 km at the equator).

To represent the topographic heterogeneity of the study region, we used the elevation data derived from the Shuttle Radar Topography Mission (SRTM) (NASA JPL, 2013) and the GTOPO30 (USG,1996) (Fick and Hijmans, 2017). This dataset contained elevation data derived from a digital elevation model (DEM) of the planet at a spatial resolution of approximately 2.5 arc-min. With this elevation data, we then calculated other metrics of interest to describe the topographic heterogeneity of the study region: aspect, slope, and terrain ruggedness. Slope and aspect were calculated as the weighted finite differences of the north-south and east-west of the 8 neighbouring pixels, which are then used to calculate the slope magnitude and aspect direction (Horn, 1981). Similarly, the Terrain Ruggedness Index (TRI), is calculated as the mean absolute elevation difference between a focal cell and its surrounding cells in a 3 × 3 neighbourhood (Wilson et al., 2007).

### Species distribution modelling

To model the potential distribution of our target species, we used a presence-only maximum entropy modelling algorithm (MaxEnt) (Phillips et al. <u>2006</u>). This approach uses a combination of presence data and background information to estimate the probability of a species occurring given a set of environmental features (Phillips et al. <u>2006</u>). MaxEnt has been extensively used in ecology and conservation, due to this, its capacity to use highly multidimensional data, and its flexibility in terms of parameterisation, make it ideal for our study’s aims (Phillips et al. <u>2006</u>).

In addition to our presence data, we generated 10,000 background points (MaxEnt default value). These points were distributed across our study area based on a spatial density kernel created by the presence records of *O. longicaudatus* (e.g Barbet-Massin et al., 2012; Jarnevich et al., 2017; Barber et al., 2022).This background sampling is meant to account for the natural asymmetry in the distribution of presence data (e.g Barber et al., 2022). In this way, we ensure that areas within the natural ranges of the species is more frequently sampled than those outside these areas. Thus, ensuring we obtain information for the whole study region without overrepresenting areas that are of no interest to the species. Once sampled, both the presence and background data were split into train and test datasets (70%-30% respectively). The train data was fed directly into the model, whereas the test data was used to calculate model accuracy and performance.

The MaxEnt algorithm relies on a family of 5 different fitting features to adjust the model responses. To generate a wider range of model responses and distribution scenarios, we randomly selected these features (Redding et al. <u>2017</u>; Albaladejo-Robles et al., 2025). Similarly, we also allow the β-multiplier to change randomly. This is a tuning parameter that controls feature expansion and regularisation strength in the model (Philips and Dudik, 2008; Merow et al., 2013). Higher β-values prevent overfitting by penalising overly complex models, thus producing simpler or smoother models. Lower β-values impose weaker regularisation, allowing more complex models to form, potentially leading to overfitting. Since we focus on the present potential distribution of the species, we set this parameter to randomly change between 1 and 6 (low to intermediate regularisation values). This way, we allow complex models not to be heavily penalised. We ran a total of 50 different MaxEnt models for *O. longicaudatus*.

Once computed, we selected the 10 models with higher reliability and performance to calculate the species potential distribution. Model selection was performed numerically, using two different metrics to measure model accuracy and reliability (Knowalik et al., 2021). The best performing models were selected using the corrected Akaike Information Criterion (AICc) (e.g Stoice and Selen, 2004), and the Boyce index (Boyce et al., 2002). The AIC is a unitless metric used to compare statistical models based on their goodness of fit and their complexity. The corrected AIC, or AICc, penalizes overly complex and overfitted models in favour of simpler but more generalizable models. For the classification of our MaxEnt models, we first ran a null model with a randomly distributed sample of pseudo-presence and absence data. This null model was used to discard all models with an AICc statistically worse than that of the null model. Once calculated, models were ranked according to their AICc scores; lower values describe better model fit, and the 10 best-performing models were retrieved.

To evaluate the performance of the ranked models, we used the continuous Boyce index. This index measures how much better a model predicts species presence when compared to a random expectation (Boyce et al., 2002). A Boyce index of 1 denotes a perfect model classification, whereas an index lower than 0 or close to -1 indicates models that perform worse than random. Values closer to 0 represent models that perform no better than a random guess (Boyce et al., 2002). For our filtering process, we discarded all models that had a Boyce index below 0.25.

### Model averaging

The resulting MaxEnt models were then used to calculate the probability ofoccurrence of *O. longicaudatus* within the study area. As a result, each MaxEnt model returns a probability surface that represents the spatial probability of occurrence of *O. longicaudatus* given a combination of fitting features and penalization parameters. This way we obtained 10 probability surfaces for *O. longicaudatus*. To produce a single estimate of species probability of occurrence, we combined the different model results into a single object by applying the arithmetic mean. This represents the compound probability of occurrence of the species based on multiple model structures and estimates.

### Supplementary Methods - R package versions

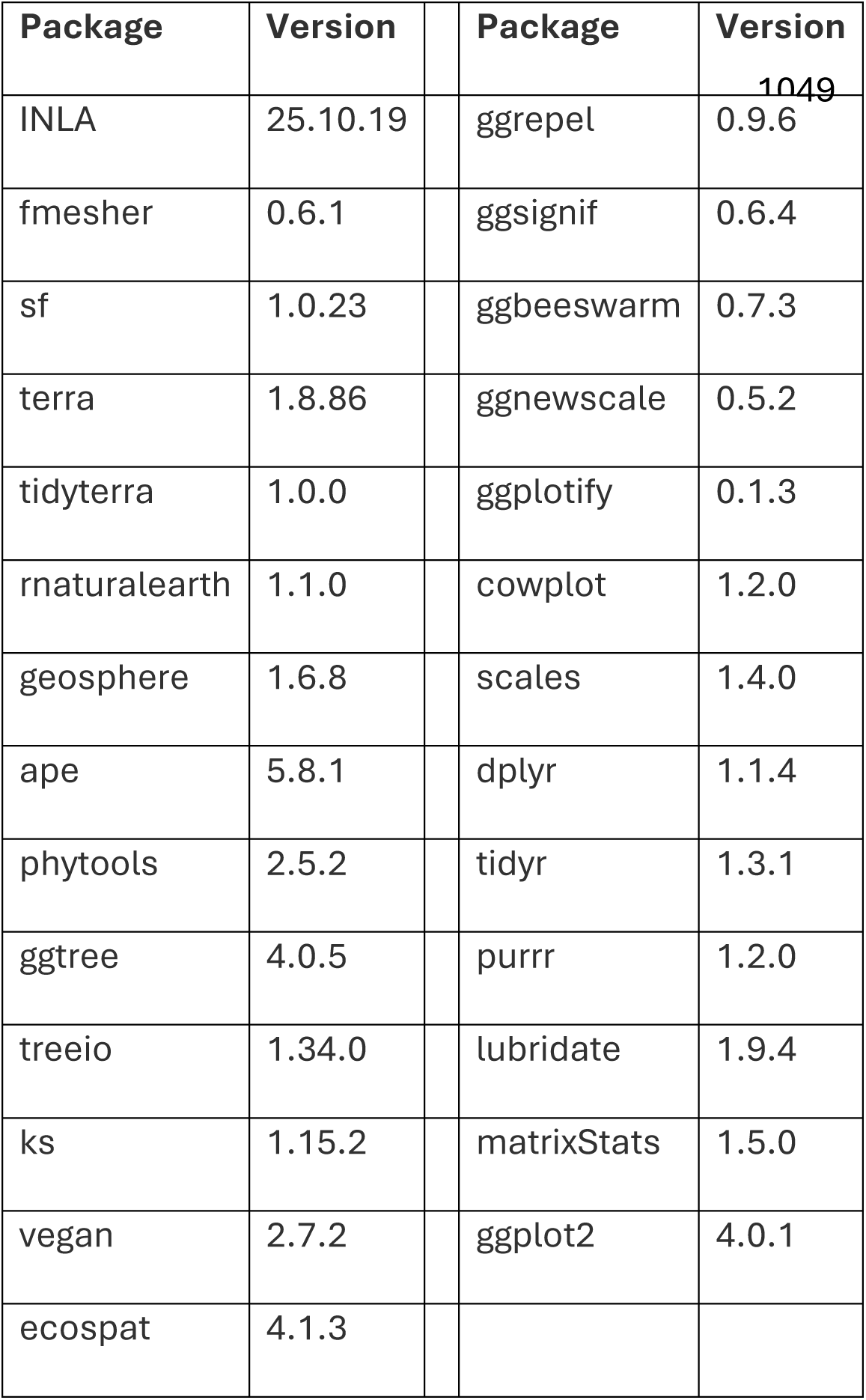

## Extended Data

**Extended Data Figure 1.**
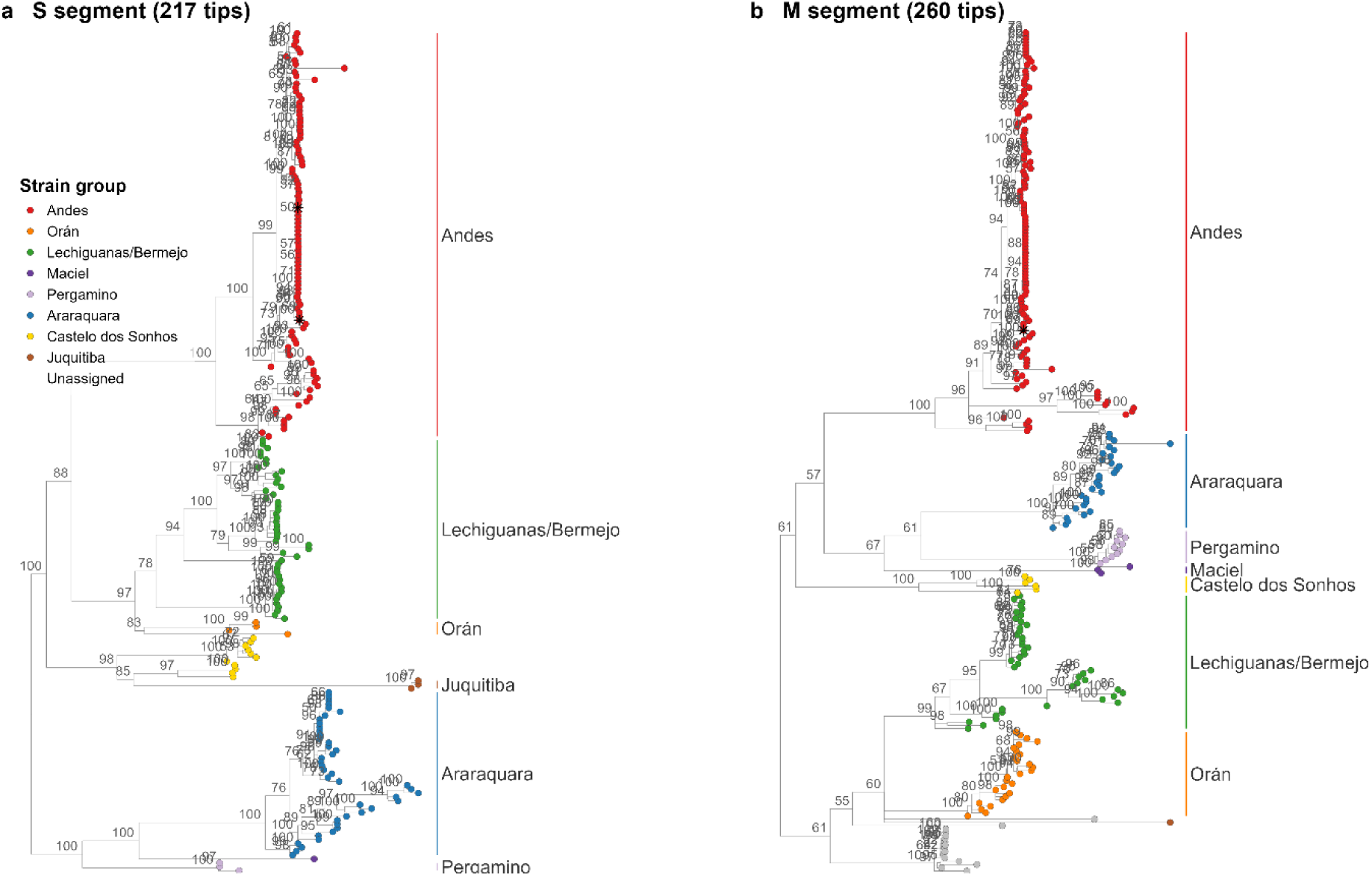
Segment phylogenies. IQ-TREE maximum-likelihood trees (S and M segments panels a and b respectively), tips coloured by tree-defined strain group, focal person-to-person strains (El Bolson 2014, Epuyen 2018-19) starred, nodes with ultrafast bootstrap < 50 collapsed, and well-supported clades labelled (see Figure 1 for support values).

**Extended Data Figure 2.**
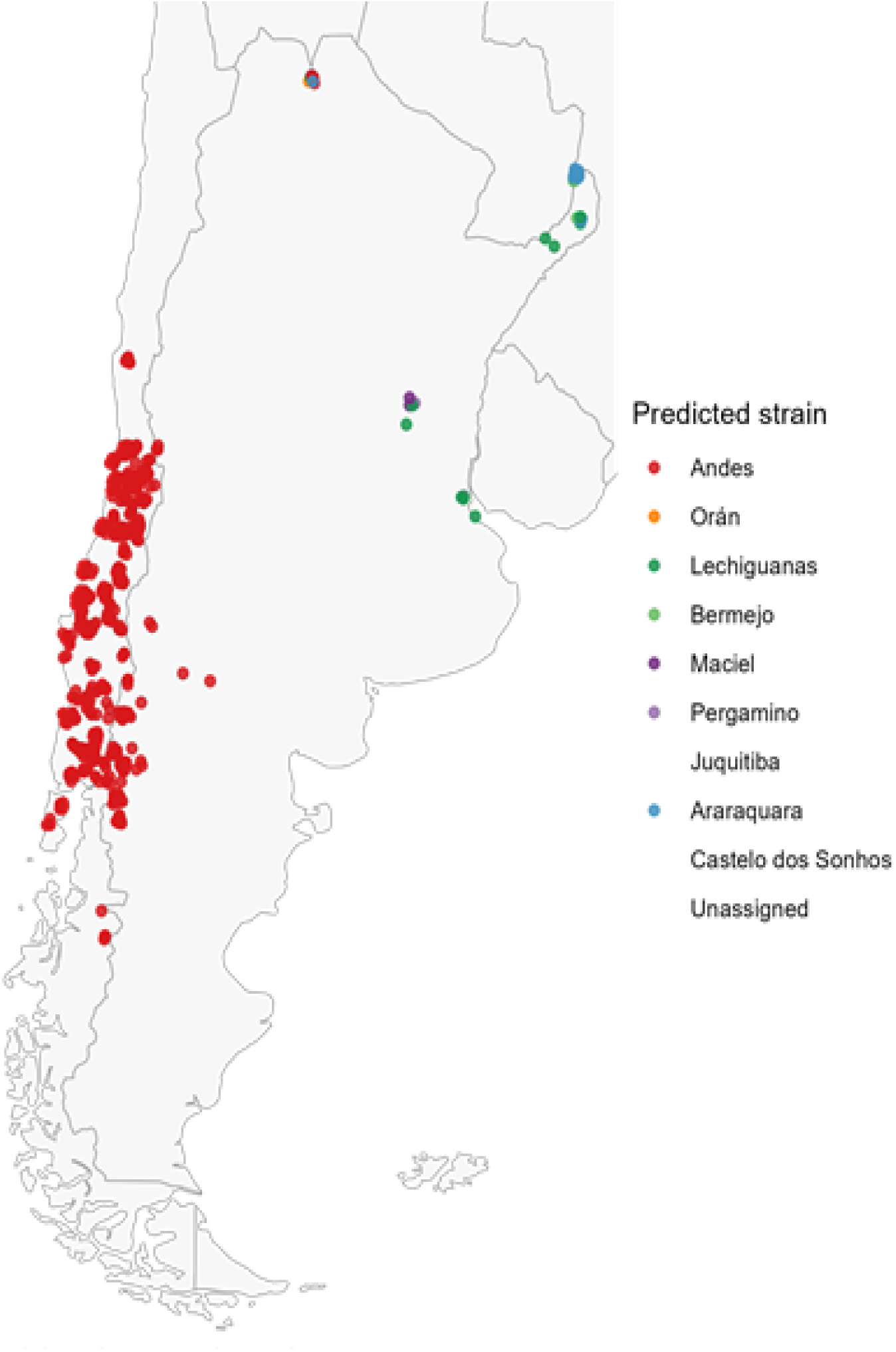
Strain attribution of serosurvey records. For each rodent serology record the most likely ANDV-complex strain is predicted from host species and locality using the same kernel typing applied to the sequences. This is used only to attribute the genus-wide serological signal to named strains (e.g. other-*Oligoryzomys* ‘ANDV’ resolves to Lechiguanas, not to the human-transmissible clade); it does not attempt to predict undiscovered viral lineages.

**Extended Data Figure 3.**
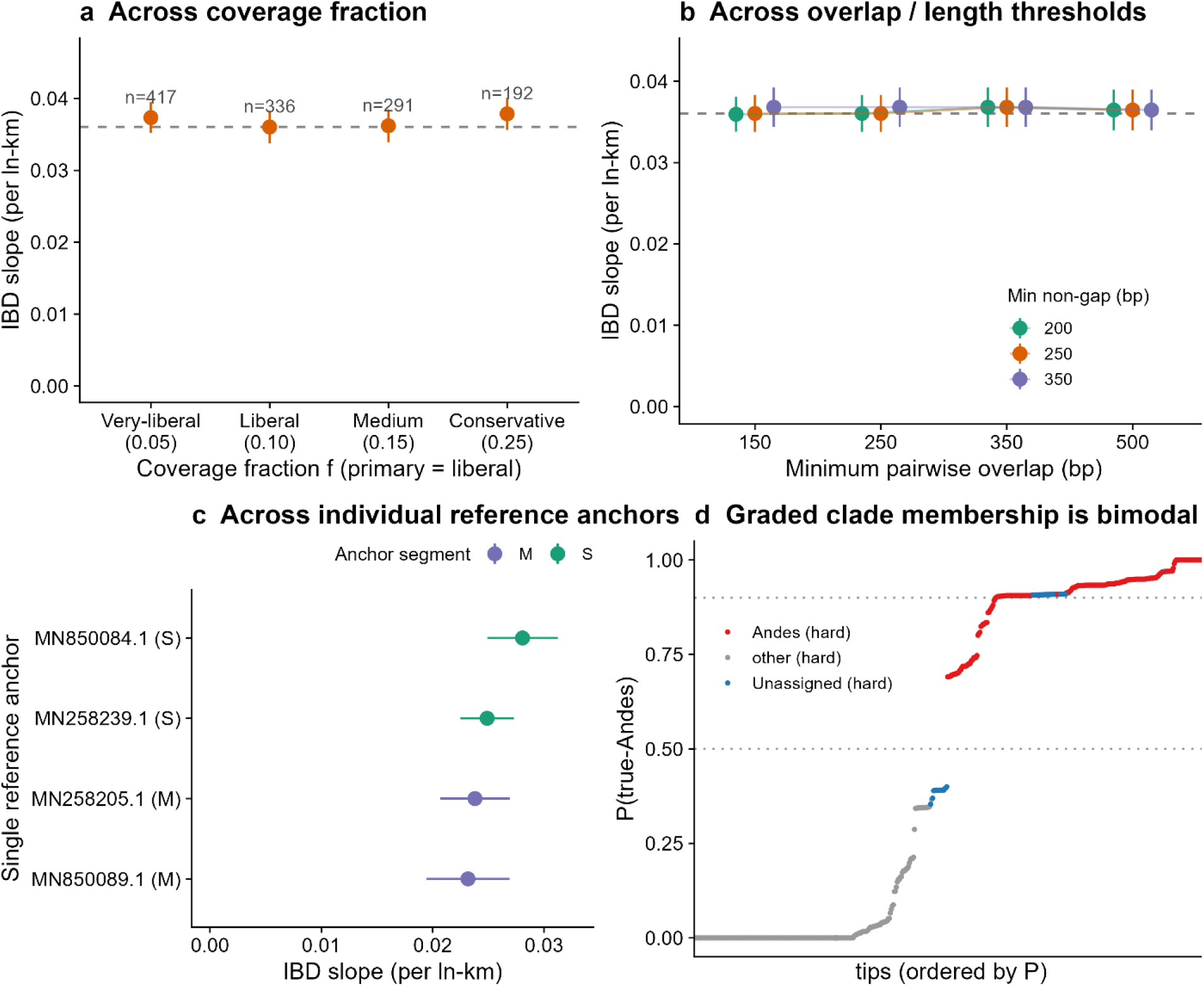
Isolation-by-distance robustness. The decay slope (∼0.036 per ln-km) is stable across (a) the coverage-fraction filter (f = 0.05-0.25; n shown per filter), (b) pairwise-overlap and minimum non-gap length thresholds, and (c) each individual reference anchor; (d) graded clade membership is bimodal.

**Extended Data Figure 4.**
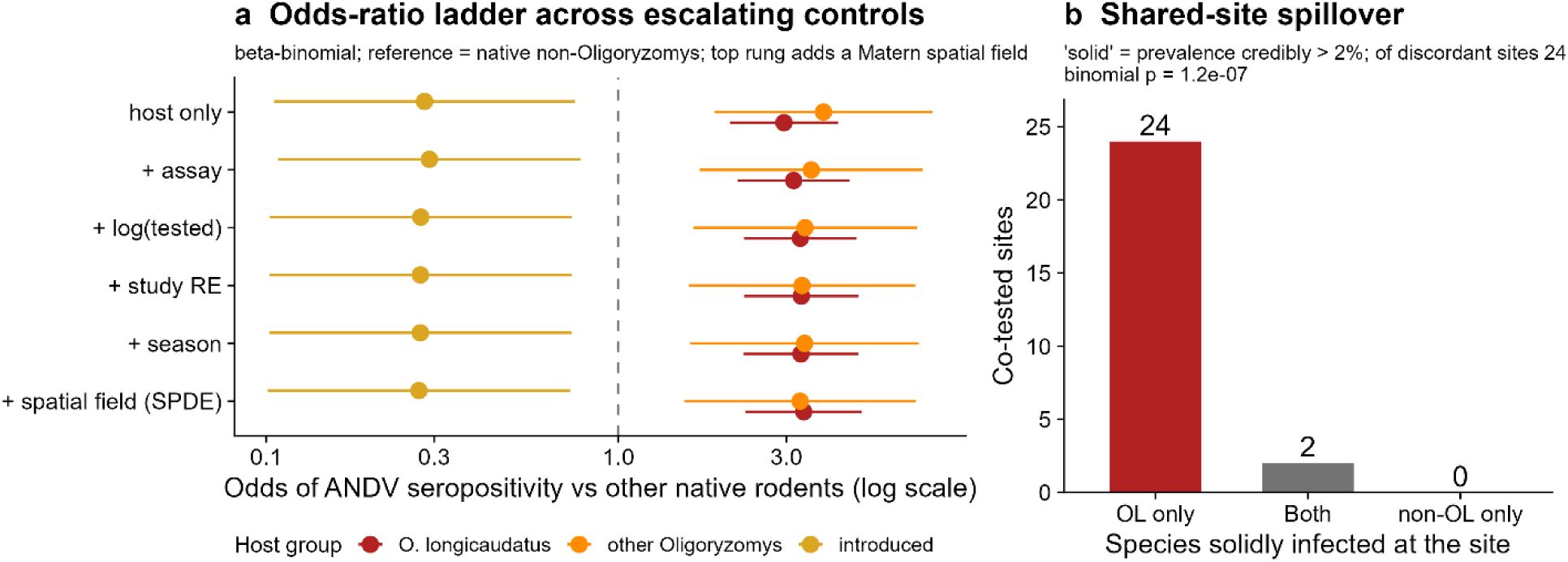
Serology robustness. (a) Odds of ANDV seropositivity in *O. longicaudatus* versus other native rodents (beta-binomial) remain ∼3x across escalating controls - assay, sampling effort, a study random effect, season, and a Matérn spatial field (the spatial-confounding test); introduced rodents are protective. (b) Shared-site spillover: co-tested sites partitioned by which species has solid evidence of infection (seroprevalence credibly > 2%): *O. longicaudatus* only (24), both (2), or non-*O. longicaudatus* only (0); discordant sites are 24:0 in *O. longicaudatus*’ favour (binomial P = 1.2e-7).

**Extended Data Figure 5.**
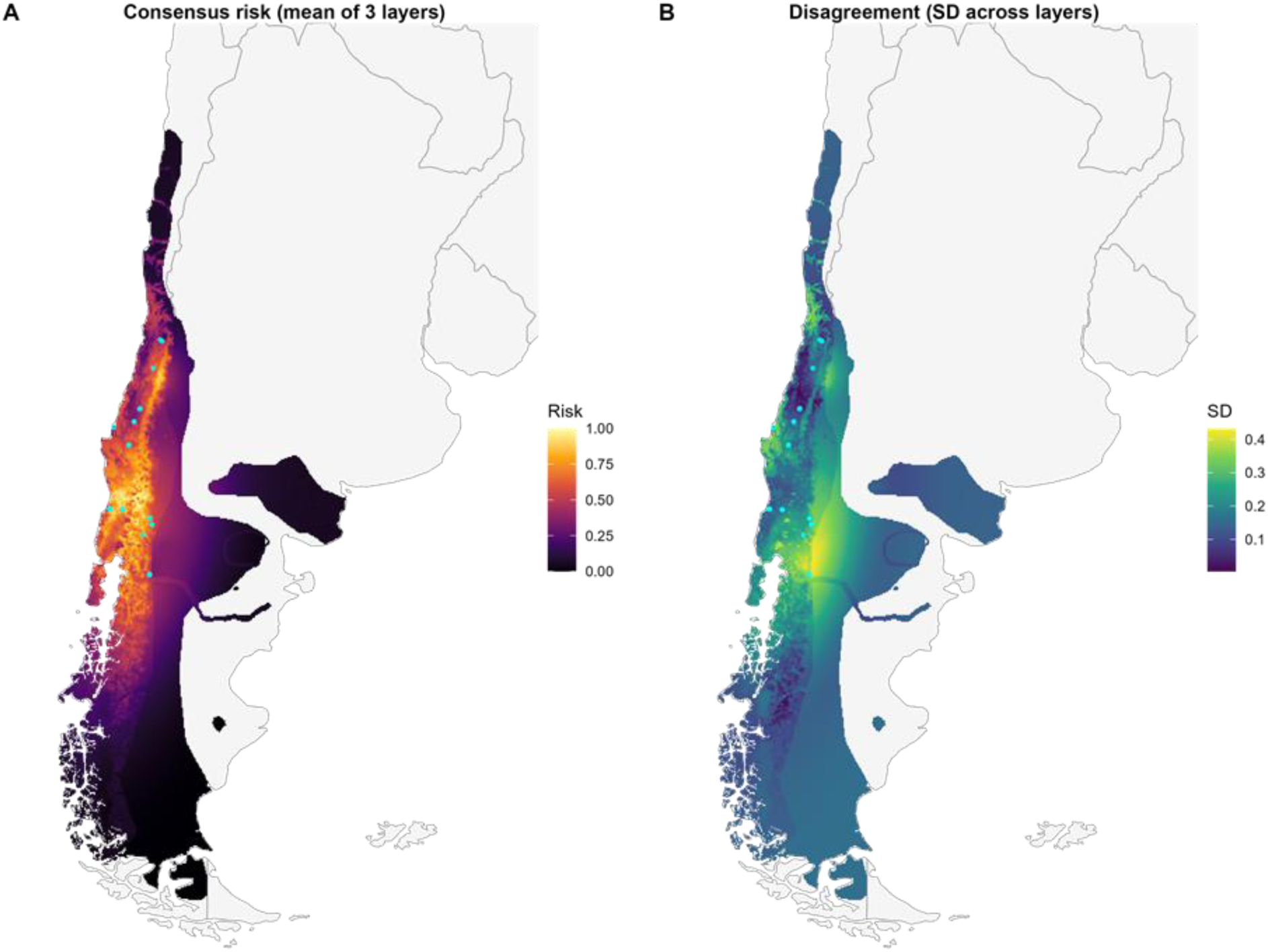
Risk-surface validation (full hold-out). (a) Consensus risk (mean of the three percentile layers) and (b) between-layer disagreement (standard deviation). ANDV human case (n = 92; cyan points) was withheld and the lineage-proximity surface refit with no human cases at all, so the risk surface was built purely from reservoir, sequence and rodent serology evidence.

**Extended Data Table 1.**
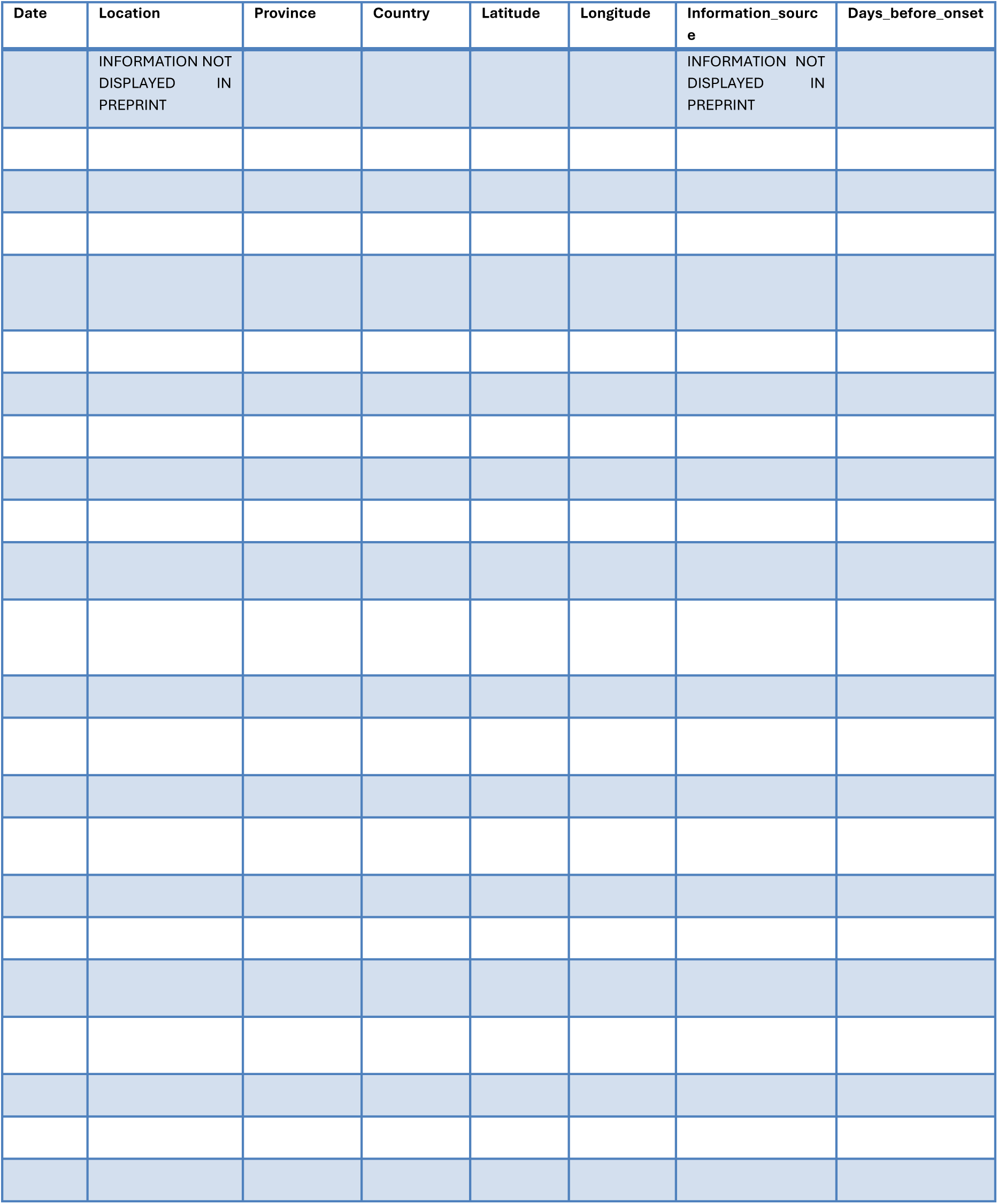
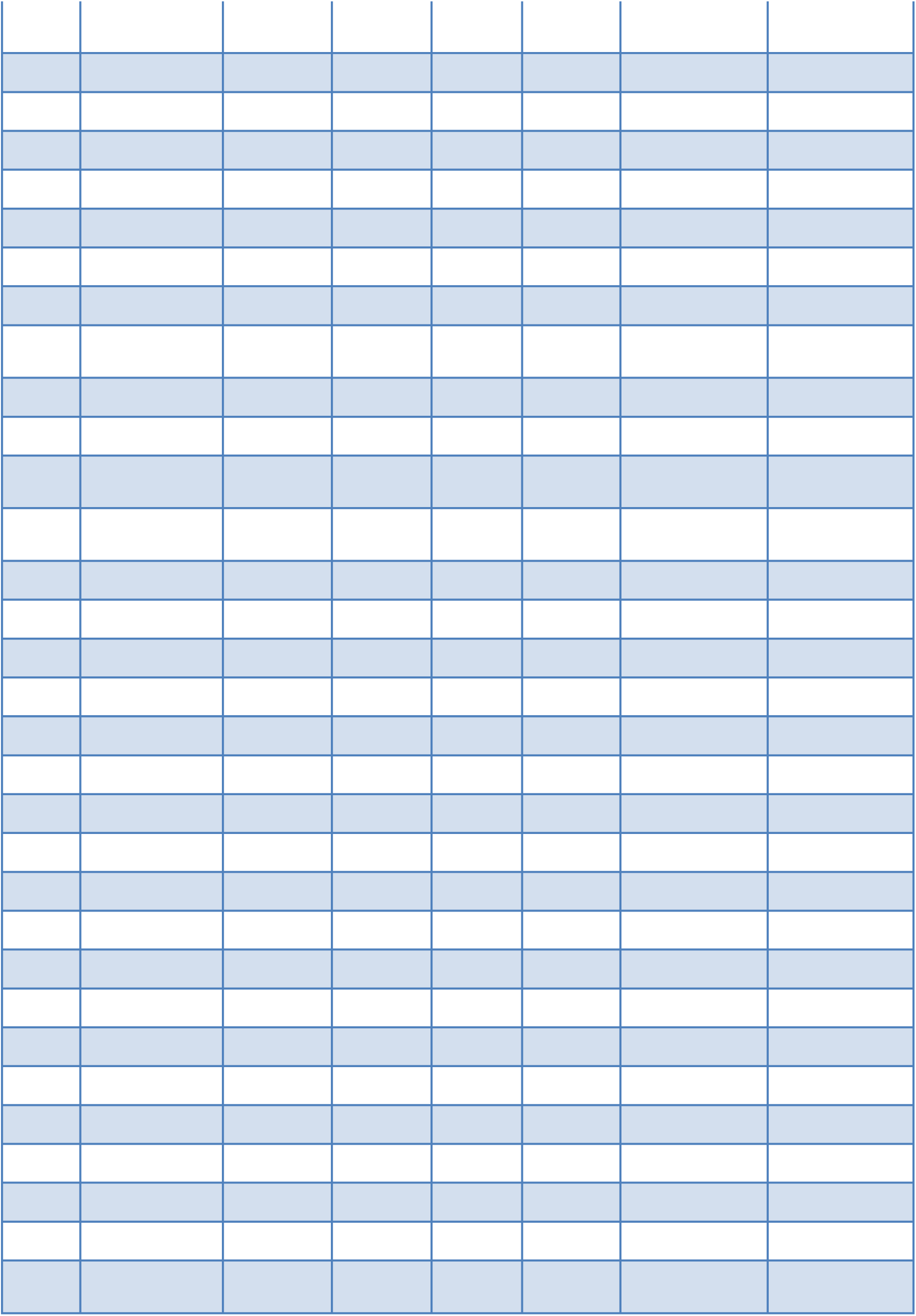
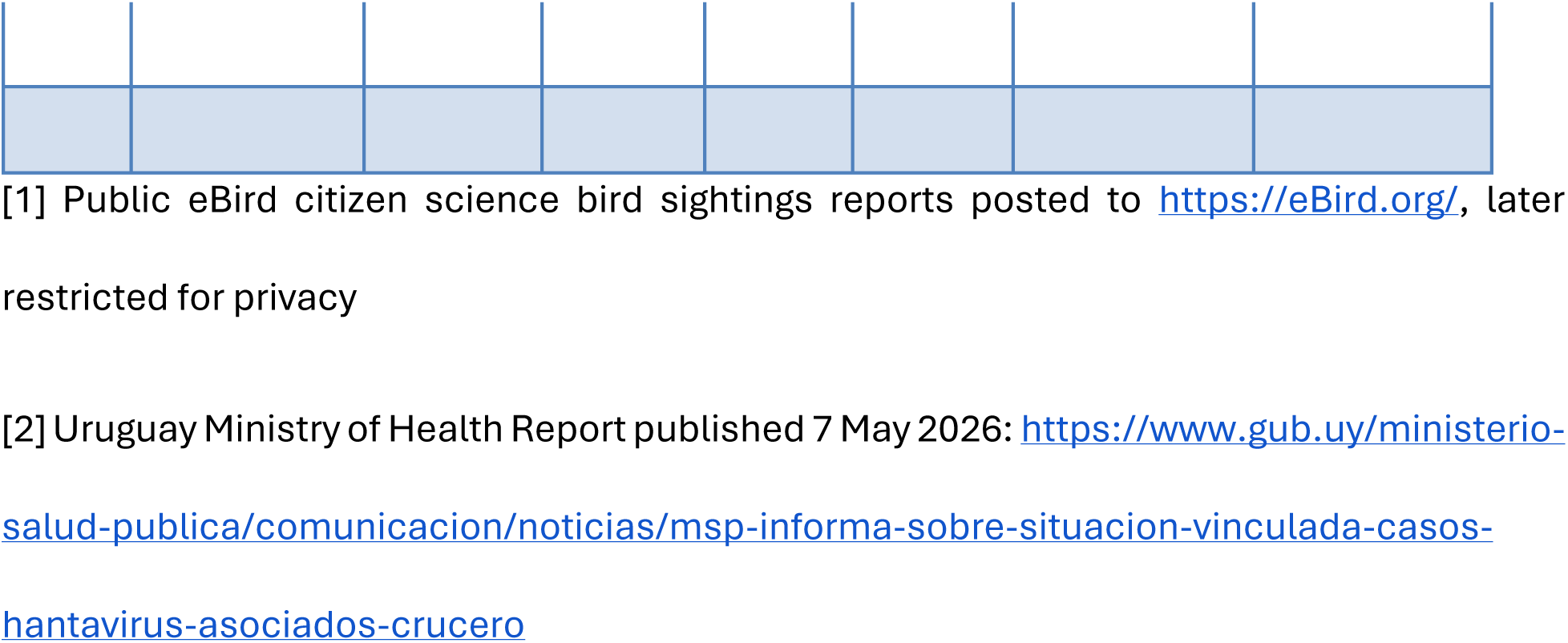
Reconstructed itinerary of the index case. Date, location, province, country, coordinates (randomly displaced 20km for privacy), information source (eBird vs media report), and days before symptom onset (3 April 2026).

